# Analysis of SARS-CoV-2 amino acid mutations in New York City Metropolitan wastewater (2020-2022) reveals multiple traits with human health implications across the genome and environment-specific distinctions

**DOI:** 10.1101/2022.07.15.22277689

**Authors:** Anand Archana, Chenghua Long, Kartik Chandran

**Affiliations:** Department of Earth & Environmental Engineering, Columbia University; 500 W. 120^th^ Street, New York, NY 10027

## Abstract

We characterize variant diversity, amino acid mutation frequency, functionality and associations with COVID-19 infections in one of the largest datasets of SARS-CoV-2 genome sequences collected from wastewater in the New York metropolitan area. Variant diversity differed within parts of the New York City sewershed and between wastewater sludge and influent samples. P314L, D614G and T3255I occurred in >95% of wastewater samples. Enhanced infectivity, transmissibility and escape from antibody neutralization were dominant traits in the wastewater. Strikingly, over 60% of the most frequently occurring mutations were found in regions other than the spike (S) protein, and nearly 50% remain uncharacterized for functional impacts warranting further investigation. We demonstrate strong correlations between P314L, D614G, T95I, G50E, G50R, G204R, R203K, G662S, P10S, P13L and mortality rates, percent positive test results, hospitalization rates and % of population fully vaccinated. The results from our study suggest that there are relatively understudied mutations in the spike protein (H655Y, T95I) and understudied mutations occurring in non-spike proteins (N, ORF1b, ORF9b and ORF9c), that are enhancing transmissibility and infectivity among human populations, warranting further investigation.

## Introduction

New York City (NYC) has been one of the epicenters of the current COVID-19 pandemic caused by the severe acute respiratory syndrome coronavirus 2 (SARS-CoV-2). COVID-19 infections have severely impaired public health, and the city faces a lasting economic toll. With the emergence of new variants (BA.2, BA.2.12 contributed to over 80% of the cases as on April 13, 2022; NY State Department of Health, 2022), control of SARS-CoV-2 continues to remain a concern owing to its demonstrated increased transmissibility, possibly affecting treatment success and vaccine effectiveness. Recently, variants BA.4 and BA.5 have been responsible for the surge in local cases. As on May 17, 2022, SARS-CoV-2 and its variants have infected 5,478,364 people in NYC (NY State Department of Health, 2022).

### 1. Viral genome sequencing: clinical samples vs. wastewater samples

Viral genome sequencing of clinical samples is an effective means of tracking SARS-CoV-2 variants in populations (Mercer & Salit, 2021) to track community spread. However, sequencing clinical samples from COVID-19 infected individuals generates information only about a single variant associated with the active infection. Additionally, the reliability of this measure depends on the proportion of clinical samples that are collected and sequenced, which may vastly under-represent the number of infected individuals. Notably, NY State Department of Health (2022) reported that <15% of clinical samples collected every month have been sequenced in 2022, and these numbers were below 11% in 2021. As of April 18, 2022, the Institute for Health Metrics and Evaluation (2022) estimated that under 10% of COVID-19 infected cases are even being detected in the United States. Therefore, health officials can no longer rely on variant detection from just clinical samples to obtain information on the relative proportion of multiple variants in the population, or to infer community spread of current and emerging variants.

Viral genome sequencing of wastewater samples can overcome some such limitations and assist in unraveling the evolution of SARS-CoV-2 or even other pathogens. Wastewater-based epidemiology (WBE) has gained substantial interest to serve as a core COVID-19 management strategy. Wastewater samples can provide a composite understanding of the overall health of a target community in a sewershed (Karthikeyan et al 2021) and wastewater data can further help public health officials track spatial and temporal changes in infections. Notably, high throughput sequencing (targeted as well as whole genome sequencing) coupled by the usage of novel bioinformatics pipelines [such as Freyja by Karthikeyan et al (2022 - preprint), LCS by Valieris et al (2020)] can efficiently document single nucleotide polymorphisms (SNPs), and amino acid mutations (Smyth et al 2020, Crits-Christoph et al 2021, Wolfe et al 2022).

### 2. Analyzing wastewater samples from a public health lens vs. a clinical lens

To date, most data on SARS-CoV-2 wastewater genome sequencing describe the early transmission of the outbreak, use data from only a few months of the pandemic, focus only on one sample type (primary sludge or influent). Previous studies (Graham et al 2021) have established the usefulness of wastewater primary sludge (samples collected from the primary sedimentation tank) for monitoring COVID-19 infections within populations. However, the effectiveness of one type over the other for SARS-CoV-2 variant surveillance remains unexplored.

Scientists have focused majority of their attention on mutations in the spike (S) protein in the SARS-CoV-2 viral RNA genome (Smyth et al 2022; Wolfe et al 2022; Heijnen et al 2021; Lee et al 2021; Graber et al 2021; Yaniv et al 2021a; Yaniv et al 2021b; Yu et al 2021) owing to its acquired ability in the neutralization of antibodies, its role in host cell entry (S protein binding with angiotensin-converting enzyme 2 (*ACE2*), and transmembrane protease serine 2 (*TMPRSS2*); Jackson et al 2022) and disease severity in human tissues (Suh et al 2022; Dai et al 2020). Such mutations have been advantageous for the evolution of SARS-CoV-2 and its variants as it spread throughout the world. For example, D614G mutation has assisted in the intermolecular association between the S1 and S2 subunits of the S protein and has been a beneficial mutation, resulting in its presence in most circulating SARS-CoV-2 variants (Jackson et al 2022). Several other variants that emerged in the pandemic were also characterized by hallmark mutations in the S protein such as L452R in Delta B.1.617.2, and N501Y, K417N and E484K in Alpha B.1.1.7, Beta B.1.351 and Gamma P.1, resulting in their partial or complete resistance to monoclonal antibodies (Starr et al 2022, Hoffmann et al 2021), justifying the much-focused appropriate scientific attention on the S protein.

Despite the unparalleled speed of vaccine development, and global mass vaccination efforts during the COVID-19 pandemic (Cascella et al 2022), enhanced transmissibility has been observed even in geographic regions where vaccine coverage has been high (North America and Europe; Gupta & Topol 2021) underscoring the need to focus on not only immune evasion but also disease spread. Focusing on the S protein might be key to tackle the pandemic in terms of development of effective treatments and specific vaccination strategies and yet might miss potentially vital information relevant to disease spread that might be present in other understudied areas of the viral genome (Alkhatib et al 2021). Notably, SARS-CoV-2 has 29 proteins – 4 structural (envelope-E and membrane-M which form the viral envelope, nucleocapsid-N which binds to the virus RNA genome, and spike-S which binds to receptors in human tissues), 16 non-structural proteins (nsp) and 9 accessory proteins (Bai et al 2022, Mariano et al 2020). RNA-dependent RNA polymerase (RdRPs) in open reading frame (ORF)1b has specific amino acids that carry out RNA-based genome replication and transcription (Shu & Gong 2016). Another example is the viral replicating enzyme 3-chymotrypsin-like cysteine protease (3CL^pro^) found in ORF1a known for its role in pathogenicity (Baloch et al 2021). However, studies regarding associated impacts of mutations in these regions remain fragmented. Therefore, from a public health perspective, viral genome sequencing of other protein regions of SARS-CoV-2, in addition to the S protein in wastewater will probably be important in predicting and mitigating disease spread in populations including viral replication, infectivity and transmissibility.

### 3. Research questions

In this study, we describe one of the largest datasets gathered from wastewater in the New York City (NYC) metropolitan area from September 2020 through February 2022. In NYC, wastewater samples (influent) were collected from Columbia University (CU) campus buildings and the North River wastewater treatment plant (WWTP) downstream of CU. Just across the Hudson River, collection of primary sludge and influent samples was done at Little Ferry WWTP in Bergen County, New Jersey (NJ). Here, we present for the first time, (a) progression of variants across target communities in NYC and NJ, (b) diversity of variants across geographies and between sludge vs. influent wastewater samples and (c) amino acid mutations in all regions of the SARS-CoV-2 viral RNA genome with associated impacts on functionality. We sought to answer the following research questions:

- What was the SARS-CoV-2 variant diversity between NYC and NJ?
- What was the SARS-CoV-2 variant diversity between wastewater influent and primary sludge samples?
- What are the most frequently occurring amino acid mutations in the wastewater?
- Is there a trend in the development of traits associated with mutations?
- Does amino acid mutation frequency correlate with intensity of expressed functionality and therefore variant emergence and progression?
- Is there an association between amino acid mutation frequency and clinical case outcomes?

## 4. Results and Discussion

### 4.1. Progression and extinction of SARS-CoV-2 variants in New York City and Bergen County

The viral composition deconvolution approach (Valieris et al 2022) revealed the maximum likelihood estimate of the relative contribution of eight WHO-labeled SARS-CoV-2 variants (Alpha B.1.1.7, Beta B.1.351, Delta B.1.617.2 and AY lineages, Gamma P.1, Iota B.1.526, Lambda C.37, Omicron BA.1, and Omicron BA.2) originating at varying time points in NYC and NJ (**Fig. 1**).

**Fig. 1.**
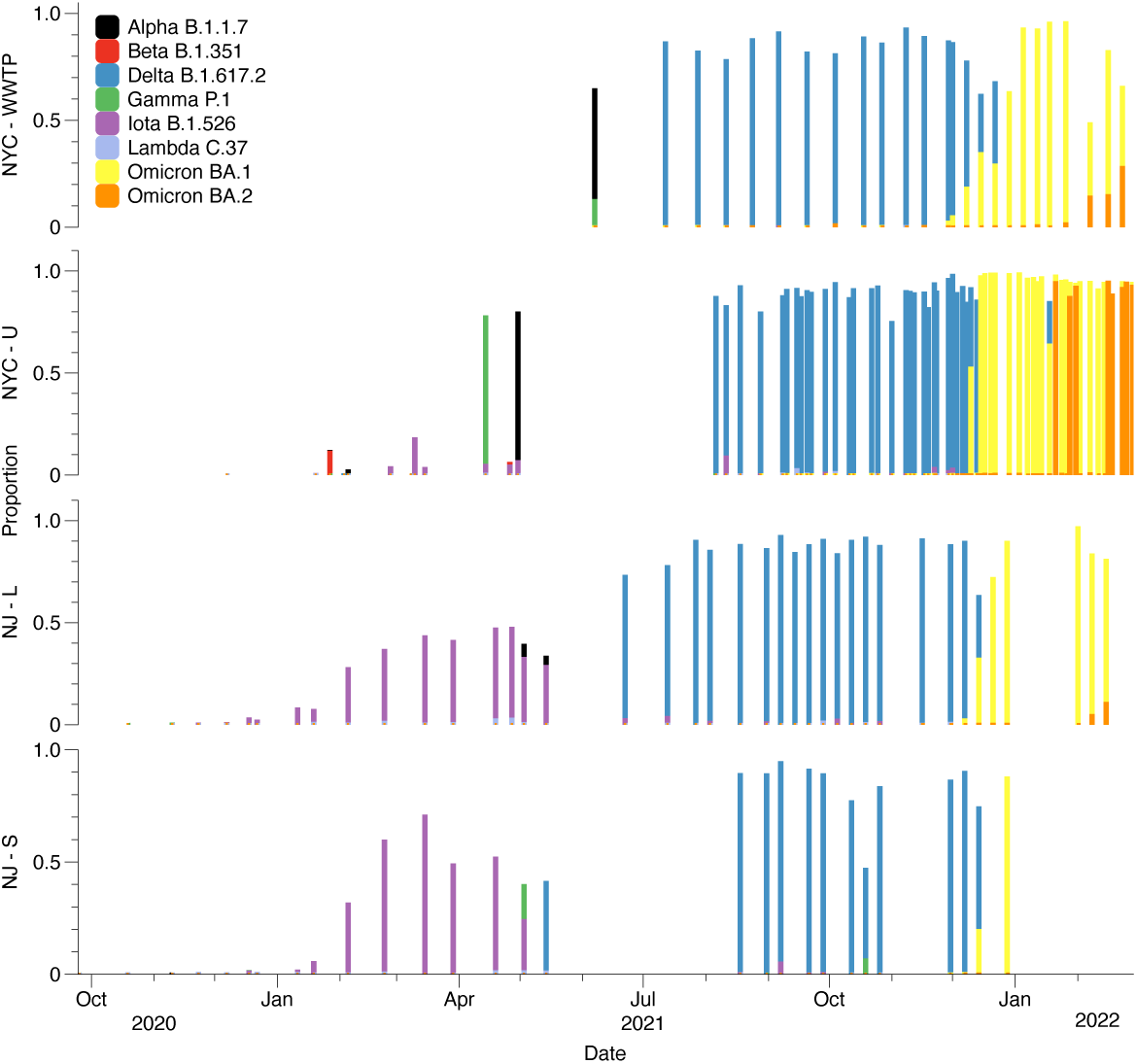
Proportion of Alpha B.1.1.7, Beta B.1.351, Delta B.1.617.2, Gamma P.1, Iota B.1.526, Lambda C.37, Omicron BA.1, and Omicron BA.2 in wastewater samples from North River WWTP (NYC-WWTP), Columbia University campus (NYC-U), Little Ferry WWTP in Bergen County (influent samples: NJ-L primary sludge samples: NJ-S) from September 2020 through February 2022.

Several drastic measures were taken to suppress the outbreak of COVID-19 infection since its emergence in the United States in 2020. The timeline of key events during the crisis (**Table 1**) provides details on the entailing damage of the pathogen on humanity and humanity’s response to it, focusing on NYC and NJ. We compared the variant proportion data to the events listed.

**Table 1.** Historical timeline of events relating to the COVID-19 pandemic in NYC and NJ (Sources: NJ spotlight news, 2022; NY Times, 2022, Investopedia 2021)

| Time | NYC | NJ |
| --- | --- | --- |
| Mar-20 | <u>Mar 1:</u> 1st confirmed case of COVID-19<br><u>Mar 7:</u> State of Emergency declared<br><u>Mar 8:</u> Guidelines issued to avoid packed buses, trains<br><u>Mar 13:</u> National Emergency declared<br><u>Mar 15:</u> Gatherings >500 to be postponed or canceled<br><u>Mar 16:</u> Public schools close<br><u>Mar 17:</u> Bars, restaurants close<br><u>Mar 22:</u> PAUSE program begins<br><u>Mar 23:</u> USNS Comfort ship arrives in NYC | <u>Mar 4:</u> 1st confirmed case of COVID-19<br><u>Mar 9:</u> State of Emergency declared<br><u>Mar 13:</u> National Emergency declared<br><u>Mar 16:</u> Casinos, racetracks, gyms, movie theaters asked to close. Indoor gatherings to be restricted to <50 people.<br><u>Mar 17:</u> Malls close<br><u>Mar 18:</u> Public schools close |
| Apr-20 | <u>Apr 6:</u> Stay-at-home order and public school closure extended to Apr 26.<br><u>Apr 15:</u> Face mask mandate in public areas<br><u>Apr 16:</u> Stay-at-home order and public school closure extended to May 16.<br><u>Apr 30:</u> Subway closures announced from 1 AM to 5 AM. | <u>Apr 6:</u> First field medical station opens in the Meadowlands<br><u>Apr 10:</u> Face mask mandate in public areas |
| May-20 | <u>May 14:</u> PAUSE order extended to May 28; State of emergency extended to June 13.<br><u>May 15:</u> drive-in theaters, landscaping, and low-risk recreational activities can reopen<br><u>May 23:</u> Gatherings of <10 permitted with social distancing | <u>May 12:</u> Non-essential retail businesses open for curbside pickup |
| Jun-20 | <u>Jun 8:</u> Phase 1 reopening begins<br><u>Jun 22:</u> Phase 2 reopening begins<br><u>Jun 24:</u> Travelers from hotspot to self-quarantine for 14 days | <u>Jun 9:</u> Stay-at-home order lifted<br><u>Jun 24:</u> Travelers from hotspot to self-quarantine for 14 days |
| Jul-20 | <u>Jul 1:</u> Beaches reopen<br><u>Jul 6:</u> Phase 3 reopening without indoor dining<br><u>Jul 19:</u> Phase 4 reopening without malls, museums, gyms, indoor dining | <u>Jul 8:</u> Universal mask mandate |
| Aug-20 | <u>Aug 4:</u> NYC health commissioner resigns | <u>Aug 13:</u> Public schools can choose how to reopen |
| Sep-20 | <u>Sep 2:</u> Gyms reopen, but indoor group workouts and pools stay closed<br><u>Sep 9:</u> Malls reopen at 50% capacity with no indoor dining. Casinos reopen at 25% capacity | <u>Sep 4:</u> restaurants, bars, salons, gyms and casinos open at 25% capacity limit |
| Oct-20 | <u>Oct 1:</u> Elementary public schools return to in-person instruction |  |
| Nov-20 | <u>Nov 19:</u> Public schools switch to all-remote | <u>Nov 10:</u> Interstate sports, restaurants, bars closed again |
| Dec-20 | <u>Dec 7:</u> Elementary public schools return to in-person instruction<br><u>Dec 15:</u> Indoor dining suspended, 1st COVID-19 vaccine administered | <u>Dec 15:</u> 1st COVID-19 vaccine administered |
| Jan-21 |  | Jan 8: Vaccine mega sites open |
| Feb-21 | Feb 11: Indoor dining reopens at 25% capacity<br>Feb 14: New Yorkers with underlying conditions become eligible for COVID-19 vaccine<br>Feb 15: Public middle schools resume in-person learning | Feb 3: Eased restrictions - NJ's indoor capacity limit increases to 35% for restaurants<br>Feb 22: Houses of worship capacity limit increases to 50% |
| Mar-21 | Mar 5: Movie theaters open<br>Mar 22: Public high schools open for in-person learning<br>Mar 30: >30 year olds become eligible for vaccine | Mar 1: Sports and entertainment venues to reopen; broader vaccine eligibility |
| Apr-21 | Apr 16: >16 year olds become eligible for vaccine<br>Apr 29: Announcement that NYC will fully reopen on July 1st |  |
| May-21 | May 12: Children aged 12-15 years eligible for vaccine | May 12: Children aged 12-15 years eligible for vaccine<br>May 28: Mask mandate ends |
| Jun-21 | Jun 1: Delta variant becomes variant of concern | Jun 4: Public health emergency ends |
| Jul-21 | Jul 1: NYC fully reopens |  |
| Aug-21 | Aug 10: NYC Governor resigns | Aug 6: Testing mandate and vaccinations for health care facilities, K-12 public schools<br>Aug 18: announcement regarding the need and availability of boosters from the Fall |
| Sep-21 | Sep 2: Broadway reopens | Sep 7: Public schools reopen |
| Oct-21 |  | Oct 18: Back-to-work reform |
| Nov-21 | Nov 3: Children aged 5 and above become eligible for vaccine<br>Nov 27: Omicron variant becomes variant of concern; State of Emergency declared | Nov 3: Children aged 5 and above become eligible for vaccine<br>Nov 29: Omicron variant becomes variant of concern; |
| Dec-21 | Dec 23: Broadway closes<br>Dec 27: CDC shortens the recommended time for isolation from 10 days for people with COVID-19 to 5 days, if asymptomatic, followed by 5 days of wearing a mask when around others. | Dec 27: CDC shortens the recommended time for isolation from 10 days for people with COVID-19 to 5 days, if asymptomatic, followed by 5 days of wearing a mask when around others. |
| Jan-22 |  | Jan 11: Public health emergency reinstated<br>Jan 19: Vaccination mandate for health care workers, correction officials and others in high-risk settings; Boosters also added to mandate |
| Feb-22 |  | Feb 2: "Pandemic" to "Endemic"<br>Feb 7: School mask mandate ends<br>Feb 28: Booster deadline for healthcare workers |
| Mar-22 | Mar 7: Students in public schools allowed to unmask; Vaccination proof required to enter gyms, restaurants, bars, lifted |  |

Alpha B.1.1.7 was first detected in the wastewater at 0.8% on 22nd December 2020 in NJ in influent samples and at a subsequent time point on 11th January 2021 in the solid samples. In NYC it was detected on 27th January 2021 (12% of pool). Since then, it rose to dominance contributing to 79.4% of the sample on 30th April 2021 in NYC and 38.9% of the sample on 3rd May 2021 in primary sludge samples from NJ (**Fig.1**). Consistent with our findings from wastewater, Alpha B.1.1.7 was noted to be dominant in NYC from January through April 2021 and faded away with the rise of the Delta B.1.617.2 variant (Duerr et al 2021). Around this time with Delta B.1.617.2’s rise, restrictions were being eased out for indoor dining, public schools began to reopen contributing to a surge in COVID-19 infections (**Table 1**).

We first detected Iota B.1.526 (also termed the New York variant) in our wastewater samples on 18th December 2020 (3% in influent; 1% in primary settled solids) in NJ and on 26th February 2021 (3%) in NYC. Subsequently it rose to dominance contributing to a maximum of 70% in March 2021. Based on clinical testing, the Iota B.1.526 variant was established in NYC around late November 2020 (Annavajhala et al 2021). Whereas the Gamma P.1 variant was introduced and established around January/February 2021 in NYC (Vasylyeva et al 2022 - preprint). We detected the Gamma P.1 variant in NYC at proportions between 0.02 to 0.05% from January through March 2021, and at 77.5% on 26th April 2021. In NJ, it accounted for 1% of the wastewater sample pool on 18th December 2020 (solid samples). Beta B.1.351 and Lambda C.37 variants were detected only in relatively small proportions in the wastewater (1%-11% and 1%-3% respectively).

Samples (B18 from NJ-S and D16 from NYC-U) collected on 15th March 2021 recorded Delta B.1.617.2 (1.3% of the pool) among other variants (B18: Alpha B.1.17 - 6.4%, Iota B.1.526 - 70.4%; D16: Iota B.1.526 - 3.2%). This is consistent with reports on the variant’s occurrence in the United States in March 2021 (Bolze et al 2022). However, it was declared a variant of concern by the United States Center for Disease Control (CDC) only on June 15th 2021, two months after it was detected in our wastewater samples.

Since then, notable surges in Delta B.1.617.2 were observed in COVID-19 infections in the populations (Insider NJ, 2021) as well as in our wastewater samples (Table S1; Fig .2; maximum 98% in NYC and 92% in NJ (influent samples)). On 27th November 2021, WHO declared the variant B.1.1.529 as a variant of concern named Omicron and on 1st December 2021, the first clinical case of B.1.1.529 was recorded in NYC (NY State Department of Health, 2022). CDC reported this variant in NYC wastewater on 4th December 2022 (CDC MMWR, 2022). In the wastewater samples we collected from NYC, we detected BA.1 at 4.75% on 1st December 2021, and at 18.2% on 8th December 2021. In NJ, we detected BA.1 at 2.2% on 7th December 2021. Subsequent increases in BA.1 (maximum 99% in NYC on 3rd January 2022 and 96% in NJ on 1st Feb 2022) are consistent with surges reported in clinical cases by CDC.

**Fig. 2.**
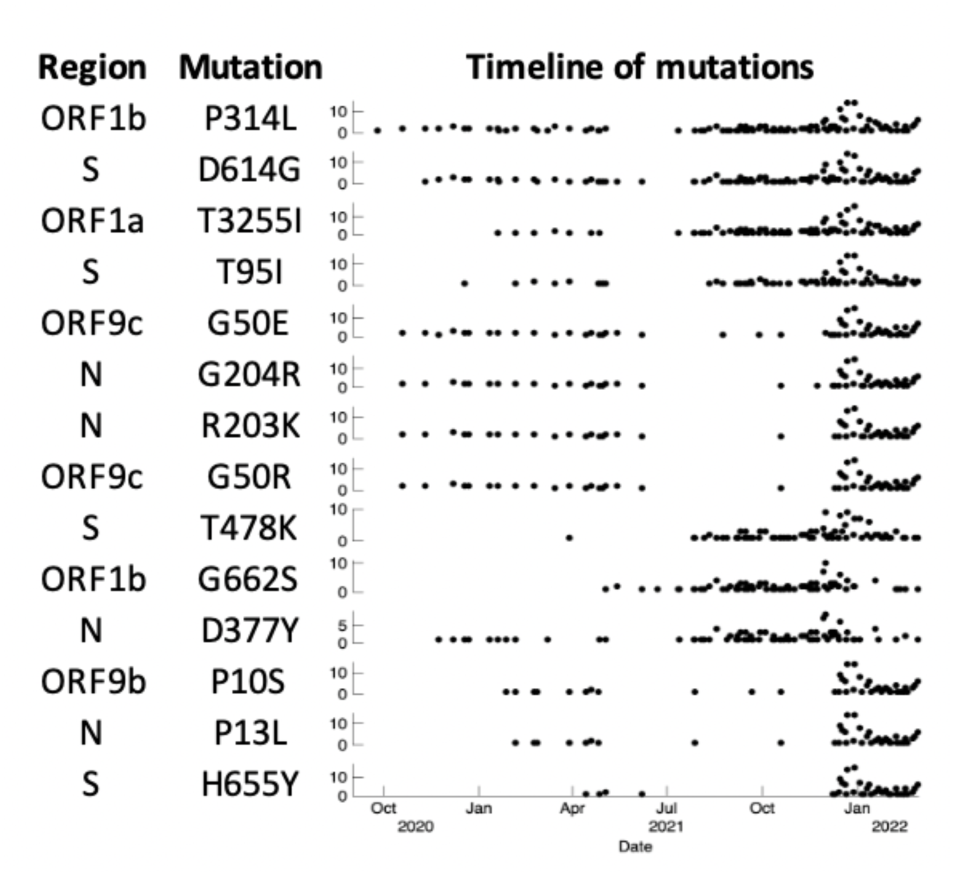
Timeline of most frequently (>50%) occurring SARS-CoV-2 amino acid mutations (in order of decreasing occurrence from top to bottom) with associated regions of occurrence within the viral RNA genome. Mutations P314L, D614G and T3255I occur in >95% of the samples.

Shortly after November 2021, concerns began to emerge concerning the sister variant of Omicron BA.1 - BA.2. The 1st clinical case of BA.2 was reported on 28th January 2022 (NBC New York, 2022). Intriguingly, we observed BA.2 at 1.1% of the wastewater samples as early as 4th October 2021 (sample H68), when the Delta variant dominated the wastewater sample at 80.5%. As the mystery unfolds concerning this variant’s origin, some studies have shown that its emergence can be traced back to as early as September or early October 2021 (Mallapaty, 2022; Peacock et al - preprint, 2022). The next occurrence of Omicron BA.2 in the wastewater samples was on 12th January 2022, two weeks ahead of the first clinical case detection. Subsequent increases (up to 95% on 16th February 2022) in variants (BA.2) from sequences obtained from reported cases were consistent with observed lineage proportions of BA.2 in the wastewater. The current study includes samples only until February 2022 and therefore does not discuss the SARS-CoV-2 variants that emerged after this time frame. However, the framework presented here can be used to discuss variants and mutations in other emerging infectious disease pathogens as well.

### 5.2. Environment specific SARS-CoV-2 variant diversity

We investigated differential patterns in SARS-CoV-2 variants at different sites including the North River WWTP in New York, NY, Little-Ferry WWTP in Bergen County, NJ and from different residential buildings on the Columbia University campus. In addition, we explored differences between wastewater influent and primary settled solid samples collected from the Little-Ferry WWTP in Bergen County, NJ.

#### 5.2.1. Distinctions in variant diversity exist within different parts of a NYC sewershed

We observed differences in variant diversity within the NYC sewershed, based on comparing samples from the CU campus buildings and the downstream North River WWTP. North River WWTP recorded a relatively higher proportion of Alpha B.1.1.7, Delta B.1.617.2, Gamma P.1, and Omicron BA.2 and samples from Columbia University campus recorded a relatively higher proportion of Beta B.1.351, Iota B.1.526, Lambda C.37 and Omicron BA.1 respectively.

North River WWTP covers a relatively larger sewershed (zip codes NY 10001, 10011, 10018, 10019, 10023, 10024, 10025, 10026, 10027, 10030, 10031, 10032, 10033, 10034, 10035, 10036, 10039, 10040) and is downstream of Columbia University campus which partially covers zip codes NY 10025 and 10027). We hypothesize that the relatively larger proportions of some variants observed in North River WWTP (Alpha B.1.1.7, Delta B.1.617.2, Gamma P.1 and Omicron BA.2) is likely to be because of the sewershed size when compared with Columbia University campus. It is likely that at the time of sampling, wastewater influent at North River WWTP had other variants that were dominating the pool (**Fig. 1**) which contributed to us not being able to detect variants such as Beta B.1.351, Iota B.1.526, and Lambda C.37. North River WWTP sewershed is home to hospitals, which might have also influenced the variant diversity information with inputs from hospitalized patients with different residential or behavioral characteristics compared to university campus residents.

Our results suggest (not unexpectedly) that the distribution of SARS-CoV-2 variants across wastewater samples (and by extension in infected communities) can be influenced by the scale of sampling. While sampling at a large sewershed scale can provide useful information on dominant variants prevailing across the community overall, a different picture could emerge by sampling sub-populations within the sewershed. Therefore, we recommend that sampling efforts be designed at appropriate resolution to provide information to specific communities of interest.

#### 5.2.2. Distinction in variant diversity exist between the liquid and solid phases of a given wastewater stream

Comparisons between the liquid influent and primary sludge from the Little-Ferry WWTP in Bergen County, NJ revealed differences in the relative proportions of the SARS-CoV-2 variants. Notably, the proportions of Alpha B.1.1.7, Beta B.1.351, Delta B.1.617.2, Lambda C.37 and Omicron BA.1 were relatively higher in influent samples and proportions of variants Gamma P.1 and Iota B.1.526 were relatively higher in primary sludge over the course of the entire monitoring program (September 2020 – February 2022). It is possible that differences in chemical composition the might have contributed to the differential detection of SARS-CoV-2 variants in the two wastewater fractions (liquid influent and primary sludge). For instance, certain humic substances are important constituents of sewage sludge (Michalska et al 2022) and are likely to deter viral replication by adsorbing onto the envelope protein and blocking cell surface entry. They have been demonstrated to have a selective inhibition effect on some naked as well as enveloped viruses such as HIV, CMV, Influenza A, and HSV-1, potentially also inhibiting SARS-CoV-2 activity (Hafez et al 2020). Another likely explanations is the presence of inhibitors that impact coverage of retrieved SARS-CoV-2 viral genome sequences in wastewater primary sludge (Baaijens et al 2021 – preprint).

Recent studies have demonstrated the sensitivity of wastewater primary sludge over influent in quantifying SARS-CoV-2 RNA concentration (Graham et al 2021) with the concentration in primary sludge being several times higher than that of influent wastewater. Additionally, the longevity of the virus on solids versus influents (studies on other human viruses including coronaviruses have demonstrated successful detection in wastewater sludge – Bibby & Peccia 2013; enveloped viruses were found to adsorb/attach to wastewater solids – Ye et al 2016, Balboa et al 2021) has been mentioned as a cause to prefer primary sludge over influent for wastewater SARS-CoV-2 RNA detection.

However, given our results demonstrating that even the variant distribution between the liquid wastewater and sludge samples could be distinct, we suggest that both wastewater phases must be considered to get a complete understanding of SARS-CoV-2 diversity present in target waste streams.

### 5.3. SARS-CoV-2 variants and their mutation characteristics

Several amino acid mutations are characteristic of the SARS-CoV-2 variants that emerged and have implications on virus attributes. Depending on the epidemiological characteristics, WHO categorized some SARS-CoV-2 variants as “variants of concern” - VOC or “variants of interest” - VOI. Here, we provide a comprehensive overview of the mutations characterizing VOCs (Omicron BA.1, BA.2 – **Fig.S1**), Delta (B.1.617.2 – **Fig.S2**), Alpha (B.1.1.7 – **Fig.S3**) and VOI Iota (B.1.526 – **Fig.S4**) as determined from the wastewater sampling program herein. Results presented in sections 5.3.1-5.3.3 refer to amino acid mutation data analysis that was conducted on samples from both NYC and NJ.

#### 5.3.1. Most frequently occurring amino acid mutations

Three amino acid mutations occurred in over 95% of the wastewater samples - ORF1b:P314L, S:D614G and ORF1a:T3255I (**Fig.2**) and eleven mutations (S:T95I, ORF9c:G50E, N:G204R, N:R203K, ORF9c:G50R, S:T478K, ORF1b:G622S, N:D377Y, ORF9b:P10S, N:P13L and S:H655Y) were found in over 50% but less than 95% of the wastewater samples. Structural proteins S and N, non-structural proteins coded by ORF1ab, and accessory proteins ORF9b and 9c were mutational ‘hotspots’ in the wastewater. However, less than 30% of the frequently occurring mutations were concentrated in the S protein. In fact, two of the 3 most frequently occurring mutations affected ORF1ab.

Mutation P314L in ORF1b occurs in the Nsp region 12, which is presumed to be part of the core replication/transcription complex and is the most conserved protein in coronaviruses (Subissi et al 2014). Thus, P314L is likely to affect RdRp activity and therefore viral RNA replication in SARS-CoV-2 (Haddad et al 2021, McAuley et al 2020). D614G has been one of the most well-studied mutations owing to its benefit to the virus for enhanced infectivity (Hossain et al 2021). Consistent with other large-scale studies (Mercatelli & Giorgi 2020) D614G and P314L were among the most prevalent mutations in the viral genome with a strong association between the two mutations (Wang et al 2021) resulting in enhanced transmissibility. Mutation T3255I is found in ORF1a in region 3CL^pro^. 3CL^pro^ is indispensable for virus replication and is essential for its life cycle (He et al 2020). Yet, to our knowledge, the associated functional impact of T3255I is currently unknown.

#### 5.3.2. Enhanced infectivity, transmissibility and escape from antibody neutralization were dominant traits observed in the SARS-CoV-2 genomes retrieved from NYC Metropolitan wastewater

Specifically, 36% of the most frequently occurring amino acid mutations (S:D614G, N:G204R, N:R203K, S:T478K and N:D377Y) were associated with enhanced infectivity (the ability of a pathogen to establish an infection), 28% (S:D614G, N:G204R, N:R203K, N:P13L) with transmission (the process by which viruses spread between hosts) and 20% (S:T95I, S:T478K, S:H655Y) with escape from antibody neutralization (**Table 2**). Importantly, the functionality of 36% of the most frequently occurring amino acid mutations (ORF1a:T3255I, ORF9c:G50E, ORF9c:G50R, ORF1b:G662S, ORF9b:P10S) remains uncharacterized.

**Table 2.**
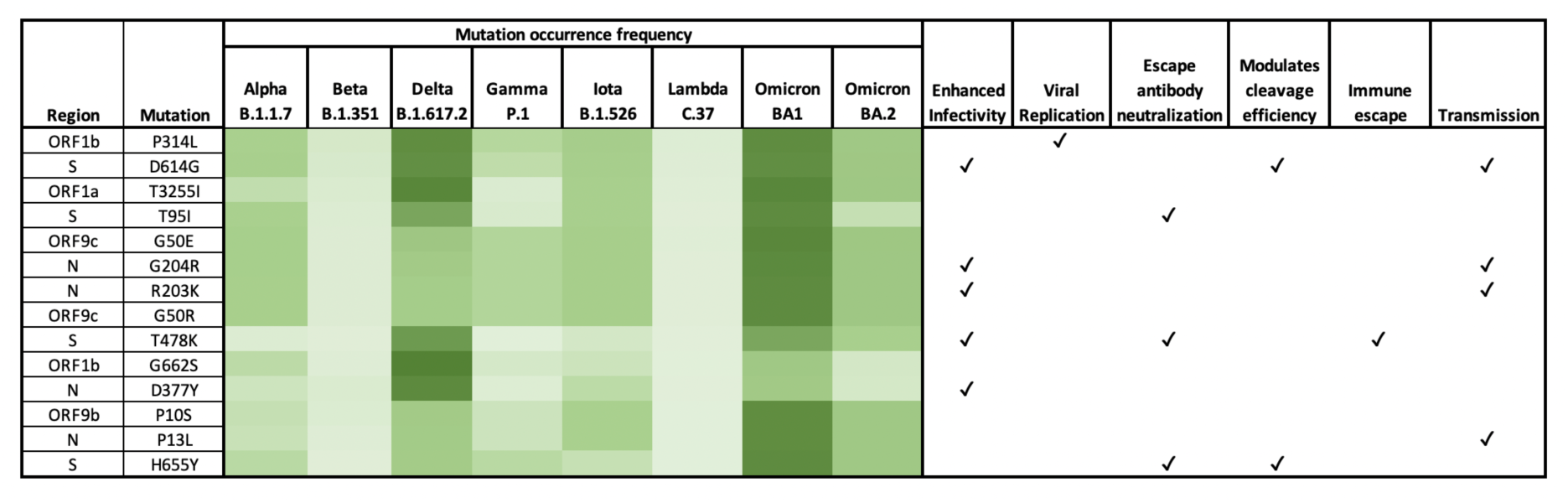
Functionality and heatmap of intensity of occurrence for the most frequently (>50%) occurring SARS-CoV-2 amino acid mutations (in order of decreasing occurrence from top to bottom) within SARS-CoV-2 Alpha B.1.1.7, Beta B.1.351, Delta B.1.617.2, Gamma P.1, Iota B.1.526, Lambda C.37, Omicron BA.1 and Omicron BA.2. Darker color indicates higher frequency of mutation occurrence.

#### 5.3.3. Enhanced infectivity and transmissibility of Omicron BA.1 and Delta

B.1.617.2 variants can be traced to specific frequently occurring amino acid mutations Table 1 shows the intensity of mutation occurrence and expressed functionality in the corresponding SARS-CoV-2 variants. The mutations appear to be distributed among different variants suggesting their important role in virus transmission and infectivity. Omicron BA.1 had transmission-associated mutations occurring at a higher intensity when compared with Delta B.1.617.2, which had infectivity-associated mutations at a higher intensity (**Table 2**).

This is consistent with reports on immune evasion which revealed that Omicron neutralization is relatively lower than that of Delta, yet its transmissibility was significantly higher (Khadia et al 2022). In the wastewater, Delta B.1.617.2 recorded a relatively high occurrence of D614G in the spike, yet mutations in the N protein - G204R, R203K and P13L, associated with enhanced transmission, were present in relatively low intensities. We observed all four mutations in relatively higher intensities in Omicron BA.1 providing evidence for the high disease spread since January 2022.

Delta B.1.617.2 recorded a relatively higher occurrence of mutations associated with enhanced infectivity (T478K and D377Y), but lower occurrence of G204R, R203K and similar intensity of D614G when compared to that of Omicron BA.1. Additionally, majority of mutations associated with immune escape, neutralization of antibodies and S protein cleavage efficiency were found in higher intensities in Omicron BA.1 than Delta B.1.617.2 except for T478K. This suggests that the mutations that contributed to the infectivity and transmissibility of Omicron BA.1 were not the same as those of Delta B.1.617.2.

#### 5.3.4. SARS-CoV-2 amino acid mutations in wastewater correlated with infection and mortality rates

We further investigated the geographic distribution of the frequently occurring amino acid mutations in NYC (North River WWTP) and NJ (Little Ferry WWTP in Bergen County) sewersheds and their correlations with mortality rates, hospitalization rates, percent positives, and % fully vaccinated (obtained from NY Department of Health for NYC and COVID-19 Info Hub for NJ from 2020 to 2022; **Table 3**, **Figs.S3-S10**). Mortality rate was calculated on a monthly basis as the average death rate per 100,000 residents, hospitalization rate refers to the average number of laboratory confirmed cases who were ever hospitalized per month, percent positive test results refer to the monthly average positive PCR laboratory results based on specimen collection date and % fully vaccinated refers to the percentage of the population that has received complete vaccine courses including Pfizer, Moderna and Janssen. Overall, the highest intensity of amino acid mutations that were significantly correlated with mortality rates were found in the following regions of the viral RNA genome: S, N, ORF1b, ORF9b, and ORF9c.

**Table 3.**
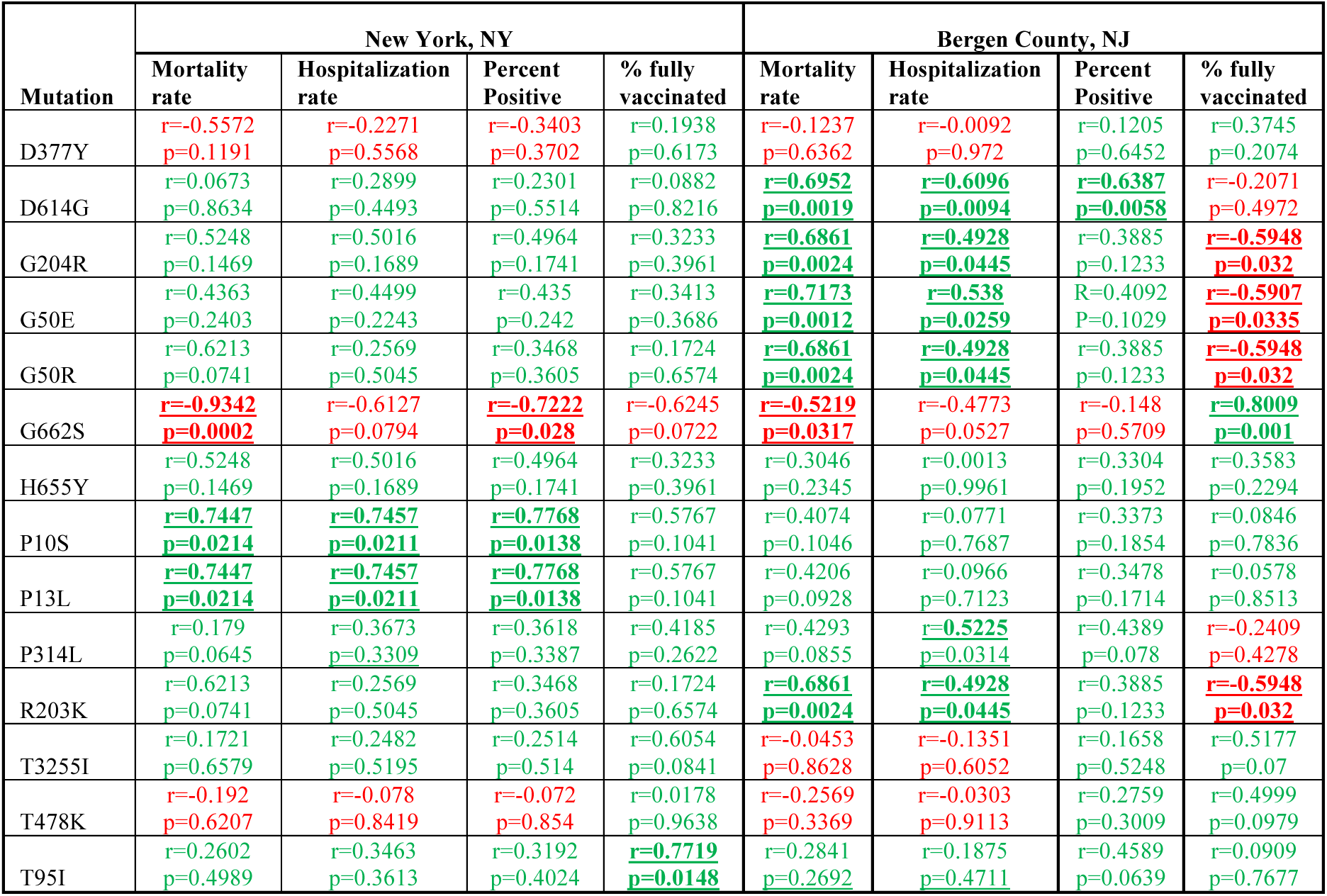
Correlation between amino acid mutations and COVID-19 related mortality rates, hospitalization rates, percent positive test results, and % fully vaccinated in NYC and NJ. Numbers in red are negative correlations. Numbers in green are positive correlations. Numbers in bold and underlined are significant correlations (*p*<0.05). Note: NYC refers to the North River WWTP and Bergen County refers to the Little-Ferry WWTP.

##### Correlation with mortality rates

In the NYC sewershed (**Table 3**), Pearson’s correlation analyses (**Fig.S3**) revealed significant positive correlations between mortality rates and ORF9b: P10S (p=0.0214; r=0.7447), N: P13L (p=0.0214; r=0.7447) and a significant negative correlation with ORF1b: G662S (p=0.0002; r=- 0.9342). Functional attributes for mutations P10S and G662S are unknown. On the other hand, we know that P13L assists in virus transmission (disease spread) (**Table 2**). Consistent with the results we saw from the wastewater, clinical case outcomes have shown that P13L was associated with fatality rates in COVID-19 infected individuals (Volz et al 2021; Eaaswarkhanth et al 2020; Magazine et al 2022; Timmers et al 2021; Gussow et al 2020; Toyoshima et al 2020). Therefore, these results demonstrate strong correlations between mortality rates and transmission-driven mutational characteristics in the NYC sewershed between September 2020 and February 2022.

At the Little Ferry WWTP in Bergen County, NJ (**Table 3**), Pearson’s correlation analyses (**Fig.S4**) revealed significant positive correlations between mortality rates and S:D614G (p=0.0019, r=0.6952), N:G204R (p=0.0024, r=0.6861), N:R203K (p=0.0024, r=0.6861), ORF9c: G50E (p=0.0012, r=0.7173), ORF9c: G50R (p=0.0024, r=0.6861)and a significant negative correlation between mortality rates and ORF1b: G662S (p=0.0317, r=-0.5219). Functional attributes of G50E, G50R and G662S are unknown. On the other hand, mutations D614G, G204R, N203K are linked with virus transmission and enhanced infectivity (**Table 2**). Consistent with our correlations in the wastewater, epidemiological observations of viral load disparities associated with D614G, G204R and N203K has shown that these mutations can increase disease severity in infected individuals, and therefore contribute to fatalities (D614G: Volz et al 2021, Eaaswarkhanth et al 2020; G203R, N203K: Gussow et al 2020; Toyoshima et al 2020). Therefore, these results demonstrate strong correlations between mortality rates and transmission-as well as infectivity-driven mutational characteristics at Bergen County between September 2020 and February 2022.

##### Correlation with hospitalization rates, and test positivity

Regarding other factors relating to infection and disease outcomes (**Table 3**), in NYC, Pearson’s correlation analyses (**Fig.S5, Fig.S6**) revealed significant positive correlations between percent positive test results and ORF9b: P10S (p=0.0138; r=0.7768), N: P13L (p=0.0138; r=0.7768) and a significant negative correlation between percent positive test results and ORF1b: G662S (p=0.0280; r=-0.7222). We also observed a significant positive correlation between hospitalization rates (Fig.S5) and ORF9b: P10S (p=0.0211; r=0.7457), N: P13L (p=0.0211; r=0.7457). Overall, the mutations that demonstrated significant correlations with mortality rates (P10S, P13L and G662S) also demonstrated significant correlations with percent positive test results and hospitalization rates in NYC.

In Little Ferry WWTP in Bergen County, NJ (**Table 3**), Pearson’s correlation (**Fig.S7, Fig.S8**) analyses revealed significant positive correlations between hospitalization rates and S:D614G (r=0.6096, p=0.0094), N:G204R (r=0.4928, p=0.0445), N:R203K (r=0.4928, p=0.0445), ORF1b: P314L (r=0.5225, p=0.0024), ORF9c: G50E (r=0.538, p=0.0259), ORF9c: G50R (r=0.4928, p=0.0445). We observed significant positive correlations between percent positive test results and D614G (r=0.6387, p=0.0058). Overall, majority of the mutations that demonstrated significant correlations with mortality also demonstrated significant correlations with percent positive test results and hospitalization rates in NYC rates (D614G, G204R, N203K, G50E, G50R). Notably, the most frequently occurring mutation in the wastewater dataset, P314L, associated with viral replication and positively correlated with hospitalization rates, was listed in the mutations associated with significant transmission events related to NYC (Gonzalez-Reiche et al 2020).

##### Correlation with % of population with complete vaccine courses

Lastly, in North River WWTP, NYC (**Table 3, Fig.S9)**, average percentage of individuals fully vaccinated was significantly positively correlated with S: T95I (p=0.0148 r=0.7719). Spike (S) mutation T95I has been shown to underlie the substantially reduced sensitivity of SARS-CoV-2 to escape from anti SARS-CoV-2 antibodies that are induced either by infection or by vaccination (Viana et al 2022) and has been associated with increased virus fitness (Obermeyer et al 2022). In fact, Magazine et al (2022) also noted the occurrence of T95I in COVID-19 infected patients that had relatively increased viral loads. Therefore, we propose that the mutation frequency of T95I increased over time owing to purifying selection pressure (Malik et al 2022).

In Little Ferry WWTP in Bergen County, NJ (**Table 3, Fig.S10**), we observed several significant negative correlations between % of population with complete vaccine course and G204R (r=- 0.5948, p=0.032), G50E(r=-0.5907, p=0.0335), G50R (r=-0.5948, p=0.032), R203K (r=-0.5948, p=0.032). It is worth pointing out that G204R, G50E, G50R and R203K were also significantly positively associated with mortality rates, hospitalization rates and percent positive test results (**Table 3**). Full vaccination against COVID-19 tends to suppress mutation frequencies (Yeh & Contrenas 2021 – preprint). Therefore, our results from wastewater testing suggest a positive correlation in declining mutation frequency with increasing vaccination coverage over the sampling period from September 2020 to February 2022.

Taken together, the results highlight key mutations with known functional attributes (P13L, P314L, T95I, D614G, G204R, R203K) and unknown functional attributes (G50E, G50R, P10S, G662S) that have likely played a vital role in prevalence and progression of the COVID-19 pandemic within the NYC Metropolitan region of focus in this study. We also note significant correlations between mutations present in the spike that are relatively understudied (such as T95I) and mutations present in other regions of the viral RNA genome (N, ORF1b, ORF9b and ORF9c) that are likely enhancing infectivity and transmissibility among human populations. In the section below, we take a closer look at these proteins and their importance.

### 5.4. Mutations in proteins S, N, ORF1b, ORF9b, ORF9c likely driving transmissibility and infectivity

The results from this study suggest that acquired mutations relating to immune escape are likely shaping the adaptation of the S protein (D614G, H655Y, T95I) and that there are understudied mutations occurring in parallel in proteins N, ORF1b, ORF9b and ORF9c, that are synergistically enhancing transmissibility among human populations.

#### Spike (S) protein

Delving deeper into the viral RNA genomic regions of SARS-CoV-2, one of the earliest S protein mutations, D614G, present in several circulating SARS-CoV-2 variants, assists in host cell entry (Korber et al 2020). Epidemiological observations of viral load disparities associated with D614G has shown that the mutation can increase disease severity in infected individuals, and therefore contribute to fatalities (Volz et al 2021; Eaaswarkhanth et al 2020). T95I positively correlated with percent positive rates (Table 3, r=0.7202) and % fully vaccinated (Table 3, r=0.7719), was also found in COVID-19 infected patients that had increased viral loads (Magazine et al 2022). Another mutation in the S protein that was found to occur frequently but was not significantly correlated with mortality rates as well as percent positive rates and hospitalization rates was H655Y. A recent study (Yamamoto et al 2021-preprint) suggests that H655Y assists in preferential pathway usage that might contribute to the pathogenesis of the variant. Intriguingly, a genome-wide automated approach for detecting viral lineages with increased fitness revealed H655Y and T95I to be the most significantly associated amino acid substitution with increased fitness ((‘fitness’ is per 5.5 days; Obermeyer et al 2022) and T95I to be one of the top five mutations for potential to spread (Maher et al 2022).

#### N protein

The N protein in SARS-CoV-2 has a vital role in viral RNA transcription and replication. We observed significant positive correlations between the most frequently occurring mutations in the N protein P13L, G204R and N203K within the wastewater and mortality rates, hospitalization rates and percent positive tests in NYC and NJ (**Table 3**). Our results are consistent with studies that have noted the occurrence of R203K, G204R, P13L and have demonstrated their association with high fatality rates (Timmers et al 2021; Gussow et al 2020; Toyoshima et al 2020) as well as infectivity (Wu et al 2021). Campbell et al (2020) predicts that P13L has a role in Cytotoxic T cell (CTL) epitope affinity and/or accessibility during infection. Additionally, the N protein is known for its involvement in several cellular processes, which makes it unique.

#### Accessory proteins ORF9b, ORF9c

Accessory proteins ORF9b and ORF9c have a key role in immune evasion (Zandi 2022; Redondo et al 2021). Specifically, the induction of Type I interferon (IFN) response is characteristic of immune defense against viral infection and ORF9b is likely to suppress this response (Han et al 2021). Recently, Obermeyer et al (2022) listed ORF9b: P10S and N: P13L among the amino acid substitutions that were most significantly correlated with increased fitness.

#### ORF1b

Within ORF1b, our most frequently occurring mutation P314L (**Table 3**, r=0.5225), associated with viral replication, had a significant positive correlation with hospitalization rates. Intriguingly, another mutation within ORF1b, G662S was significantly negatively correlated with mortality rates in both NYC and NJ (**Table 3**, r=-0.9342 and r=-0.5219 respectively). We also observed a significant positive correlation between G662S (**Table 3**, r=0.8009) with % of population with complete vaccine courses. This suggests that there was a decrease in the frequency of G662S as vaccination coverage increased. However, the significance of these correlations is unknown. To date, there is fragmented evidence concerning the occurrence of G662S in COVID-19 infected patients. Notably, Umair et al 2021 recorded the occurrence of G662S in SARS-CoV-2 infected individuals during a third wave in Pakistan. Lucas et al 2021 noted the presence of G662S in individuals who were previously infected (with Delta B.1.617.2) or uninfected and had received mRNA vaccinations to SARS-CoV-2. Rosenthal et al 2020 – preprint also noted the occurrence of G662S in clinical specimens.

Through this study, we highlight the most frequently occurring amino acid mutations within a long-term wastewater monitoring dataset in NYC and NJ and demonstrate strong correlations between understudied mutations in the spike and several mutations in non-spike regions of the viral RNA genome with COVID-19 infection outcomes. Our results suggest that within the S protein, H655Y and T95I are key mutations that have received relatively less research attention when compared with D614G. However, their associated impacts on infection levels and transmission remains yet to be characterized. Additionally, there is a significant paucity of information concerning the role of mutations in non-spike regions - N, ORF9b, ORF9c, ORF1b on the emergence, and progression of SARS-CoV-2 variants, therefore warranting further investigation.

## 6. Conclusion

With the administration of vaccinations and better treatment options, SARS-CoV-2 variants have adapted to human cells, resulting in an increase in case levels (infections and hospitalizations) yet decreased infection-related deaths. Taken together, the observed ongoing mutation progression might need variant-mutation specific vaccines and is likely to be associated with variable public health response needs. Overall, our observations are based entirely on wastewater data, and we recognize that follow up with clinical data will be crucial. However, our evidence may be an asset, perhaps even a leading indicator of future adaptations within the SARS-CoV-2 viral RNA genome that will be helpful in containing the virus.

## Data Availability

All data produced in the present study are available upon reasonable request to the authors

## Funding

This research is supported by the National Science Foundation (CBET 2026599) and the National Institutes of Health (U01DA053949).

Assistance and coordination relating to sample collection and transport by Bergen County Utilities Authority, AECOM, New York City Department of Environmental Protection and Columbia University Facilities is gratefully acknowledged.

## Supplementary Materials

### Materials and Methods

#### 1. Virus concentrated from wastewater sample

The virus concentration protocol was based on the procedure as previously described^1^. Initially, 12.5 mL of glycine buffer (0.05 M glycine, 3% beef extract, pH = 9.6) was added to 100 mL of wastewater sample, and then shake vigorously. The sample was centrifuged at 8,000×g for 30 minutes at 4℃, and the supernatant was filtered through 1 µm and 0.45 µm polyethersulfone (PES) membrane (Tisch Scientific, OH). PEG/NaCl stock solution (400 g/L PEG 8000, 87.5 g/L NaCl, Thermo Fisher Scientific, MA) was prepared and added to the filtered supernatant to achieve final concentrations of 80 g/L PEG 8000 and 17.5 g/L NaCl. Viruses were precipitated from the supernatant by incubation during agitation (100 rpm) overnight at 4℃, followed by centrifugation for 90 minutes at 4℃. The resulting viral-containing pellet was suspended with 0.6 mL phosphate buffer saline (PBS, Thermo Fisher Scientific, MA). After December 8^th^ 2021, campus samples were processed using Concentrating Pipette Select (InnovaPrep LLC, MO). Briefly, 1 mL 5% Tween 20 solution was added to 100 mL of wastewater sample, and then the sample was centrifuged at 8,000×g for 30 minutes at 4℃. Supernatant was filtered through a 0.05 um PS Hollow Fiber concentrating pipette tip (InnovaPrep LLC, MO), and the retentate was eluted twice with Elution Fluid-PBS (InnovaPrep LLC, MO).

#### 2. Viral RNA extraction and RT-qPCR

Viral RNA was extracted using QIAcube with QIAamp Viral RNA Mini Kit (Qiagen, Hilden, Germany). SARS-CoV-2 RNA concentrations were quantified by one-step RT-qPCR on a CFX384 Touch Real-Time PCR Detection System (Bio-Rad, CA) using the U.S. Centers for Disease Control and Prevention (CDC) N1 and N2 primers sets^2^. The RT-qPCR amplifications were performed in 20 µL reactions which contained 8.5 µL nuclease free water (Qiagen, Hilden, Germany), 1.5 µL N1 or N2 primer-probe mixes (2019-nCoV RUO Kit, Integrated DNA Technologies, IA), 5 µL TaqPath 1-Step RT-qPCR Master Mix (Thermo Fisher Scientific, MA), and 5 µL RNA samples. Thermal cycling reactions were carried out as follows: 25℃ for 2 min, 50℃ for 15 min, 95℃ for 2 min for reverse transcription, followed by 45 cycles of 95℃ for 3 s, 55℃ for 30s. Standard curve was generated via serial 10-fold dilutions of plasmids contain the complete nucleocapsid gene from SARS-CoV-2 (Integrated DNA Technologies, IA). The limit of quantification is the lowest copies in standard curve which is 10 copies/reaction.

#### 3. Next generation sequencing, bioinformatics and data analysis

RNA (10 µL) was converted to cDNA with the Superscript IV first-strand synthesis system (Invitrogen Life Technologies, CA) using the random primers according to manufacturer’s instruction, with a modification of increasing the RT incubation step from 10 minutes to 50 minutes. 10 µL of the resulting cDNA was used to prepare library using Swift Normalase Amplicon SARS-CoV-2 NGS Panels (SNAP) following manufacturer’s instruction. Libraires were sent to GENEWIZ, Inc. to check the libraires quality and sequencing. Briefly, libraries validation and quantification were done using TapeStation (Agilent Technologies, CA), and Qubit 2.0 Fluorometer, respectively. Libraries were loaded and 2×150 paired-end (PE) sequencing using an Illumina HiSeq platform according to manufacturer’s instructions (Illumina, CA). Sequencing reads were processed using Swift Biosciences SARS-CoV-2 analysis pipeline (https://github.com/swiftbiosciences/sarscov2analysis_docker). Briefly, adapters and low-quality bases were trimmed with Trimmomatic v0.36^3^. Sequencing reads were mapped to SARS-CoV-2 reference genome Wuhan-Hu-1 (NC_045512.2) using BWA-MEM v0.7.17-r1188^4^. Mapped reads were soft-clipped to remove SNAP tiled primer sequences using Swift Bioscience primerclip v0.3.8 (https://github.com/swiftbiosciences/primerclip). Coverage was calculated using bedtools v2.27.1^5^. The Genome Analysis Toolkit (GATK) v4.1.8.1 was used to call mutations^6^. Sequences with less than 80% genome coverage at read depth 10x were discarded from further analysis. The relative proportions of selected SARS-CoV-2 lineages were calculated according to a mixture model for determining SARS-CoV-2 variant composition in pooled samples^7^. Libraries were prepared using NEBNext ARTIC SARS-CoV-2 Companion Kit (New England Biolabs) with the V3 ARTIC primers.

#### 4. Statistical and Data Analysis

For the North River WWTP sewershed within NYC, we obtained data for mortality rates, hospitalization rates, percent positive test results and % of population fully vaccinated for specific zip codes NY 10001, 10011, 10018, 10019, 10023, 10024, 10025, 10026, 10027, 10030, 10031, 10032, 10033, 10034, 10035, 10036, 10039, 10040. Little Ferry WWTP serves roughly 50% of the population of Bergen County (Personal communication from Mr. Dominic Disalvo, Director of Engineering). Therefore, we correlated results from wastewater samples obtained from Little Ferry with health data from BC as a whole. Similar analysis was not possible for CU since zip-code related data includes both CU and non-CU populations. Pearson’s correlation analysis was done using JMP 16.2.0, heat maps were made using Microsoft Excel and data visualization was done using Datagraph 5.0.

## Figures & Tables

**Fig. S1.**
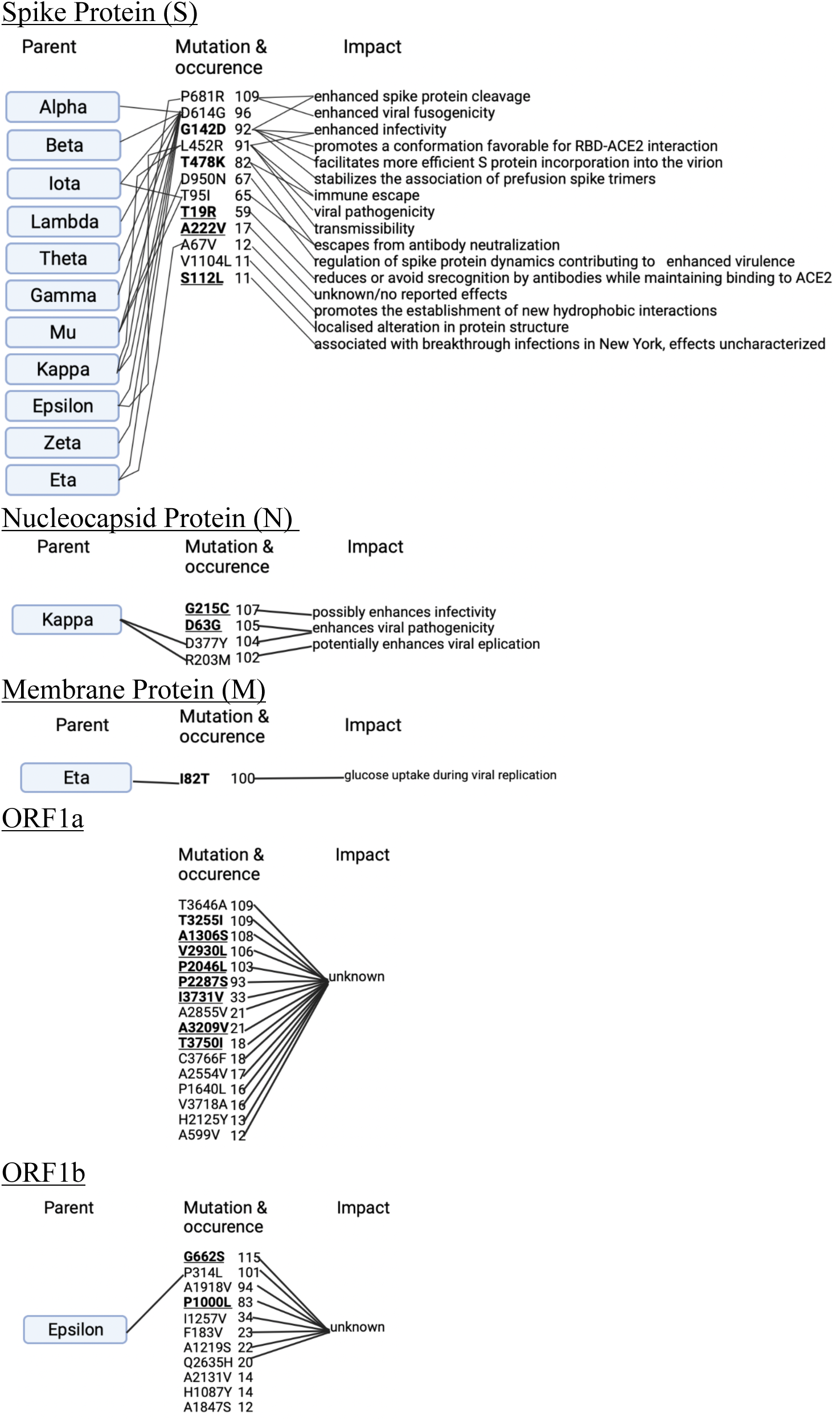
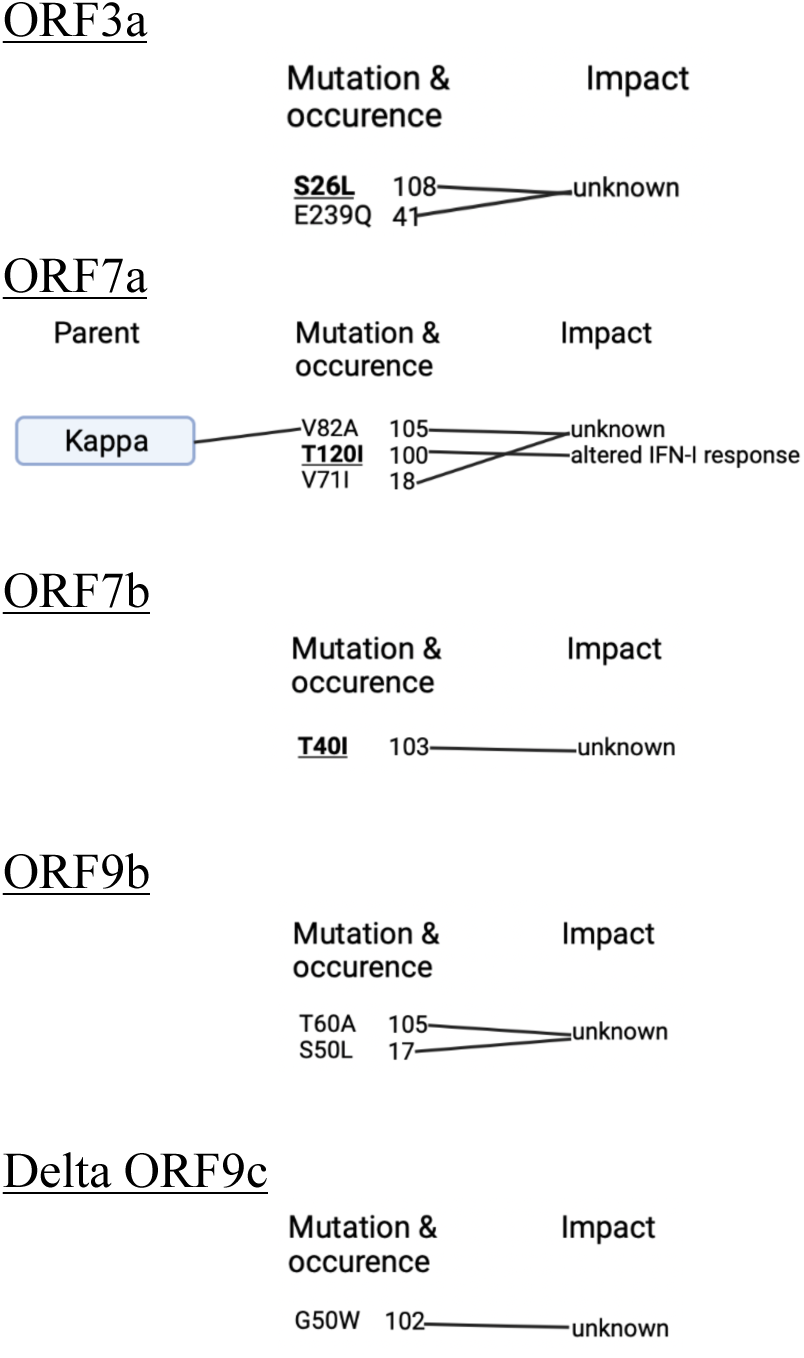
Amino acid mutations (occurring in at least 90% of the samples) in the wastewater for Delta B.1.617.2, parental origins and associated impacts. Mutations highlighted in **<u>bold and</u> <u>underlined</u>** are unique to this variant; mutations highlighted in **bold only** were first observed in this variant and are subsequently observed in other descendants such as Omicron BA.1 and BA.2; corresponding numbers indicate the frequency; mutations in parentheses indicate mutations that are known to occur together.

**Fig. S2a.**
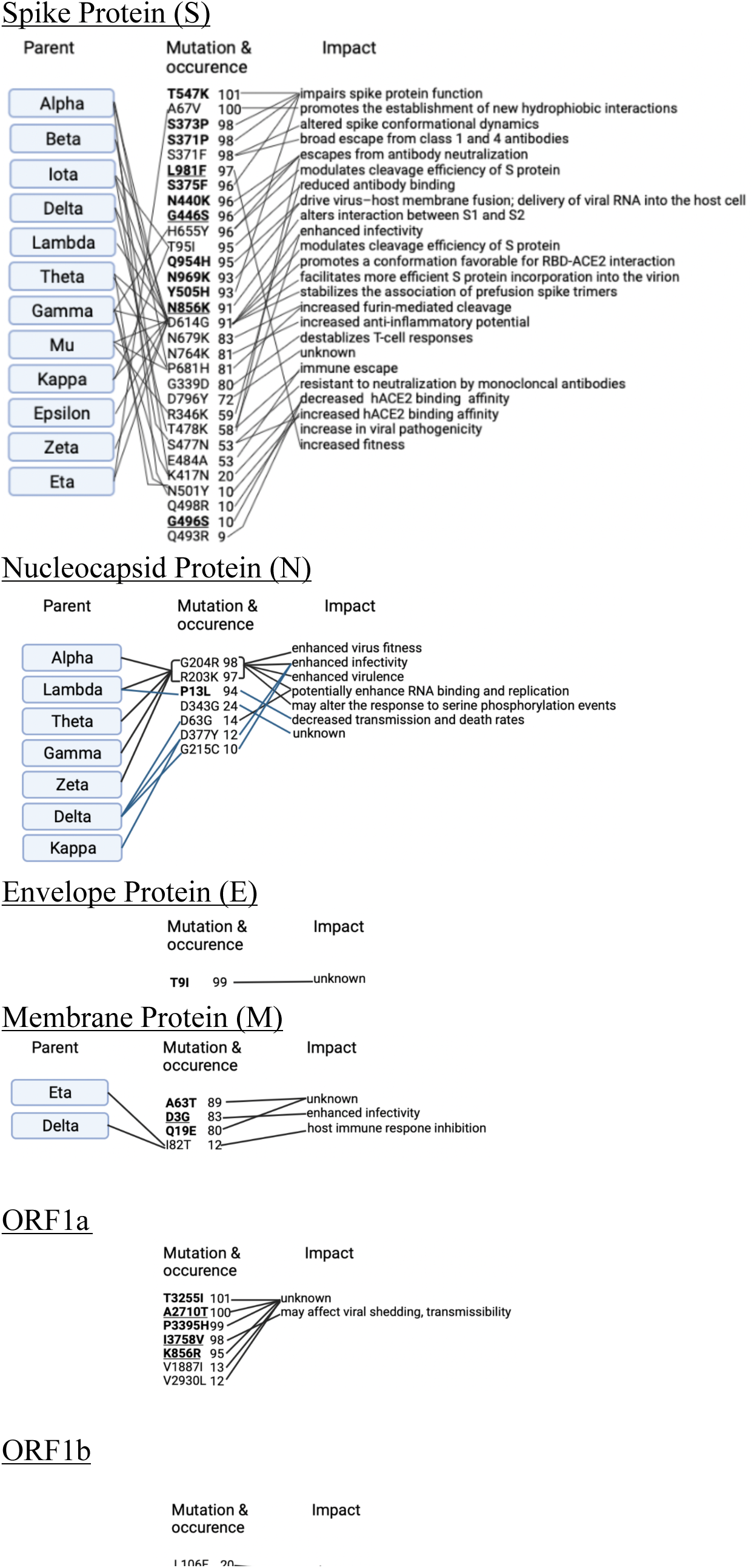
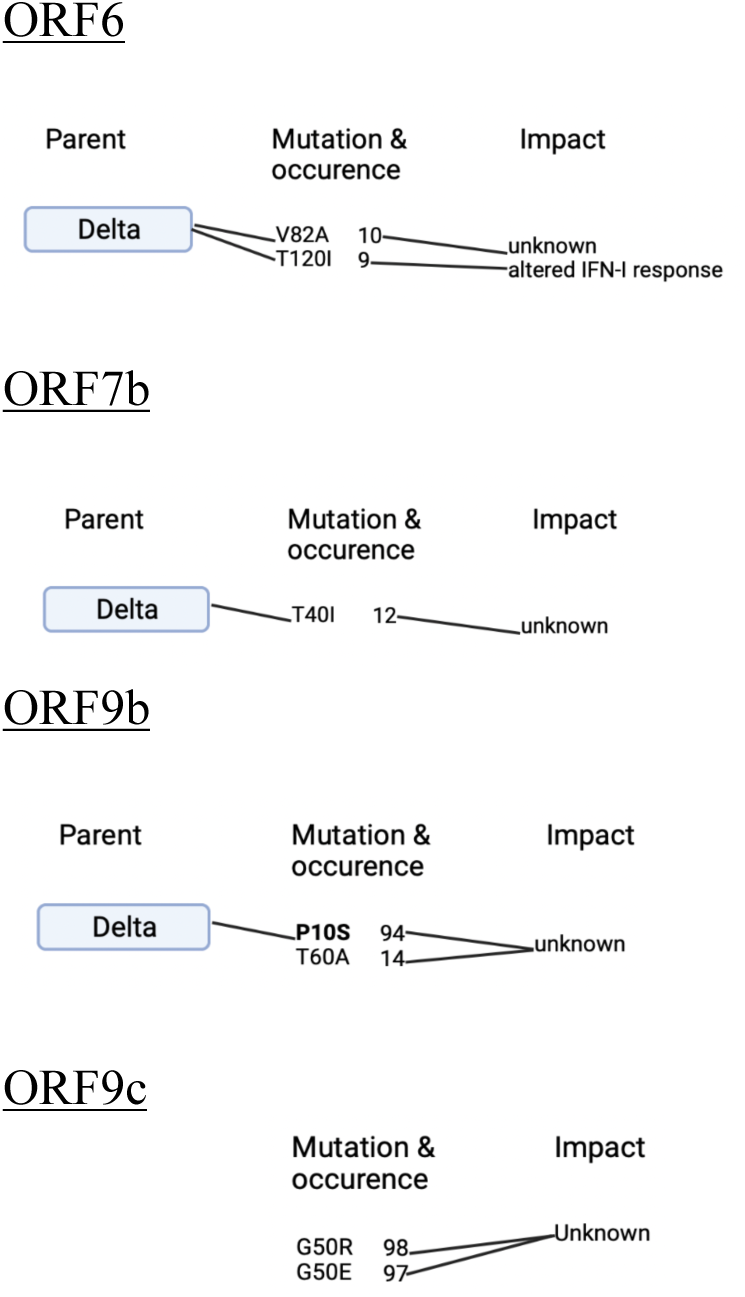
Amino acid mutations (occurring in at least 90% of the samples) in the wastewater for Omicron BA.1, parental origins and associated impacts. Mutations highlighted in **<u>bold and</u> <u>underlined</u>** are unique to this variant; mutations highlighted in **bold only** were first observed in this variant and are subsequently observed in descendants Omicron BA.2, BA.2.12.1, BA.4 and BA.5; corresponding numbers indicate the frequency; mutations in parentheses indicate mutations that are known to occur together.

**Fig. S2b.**
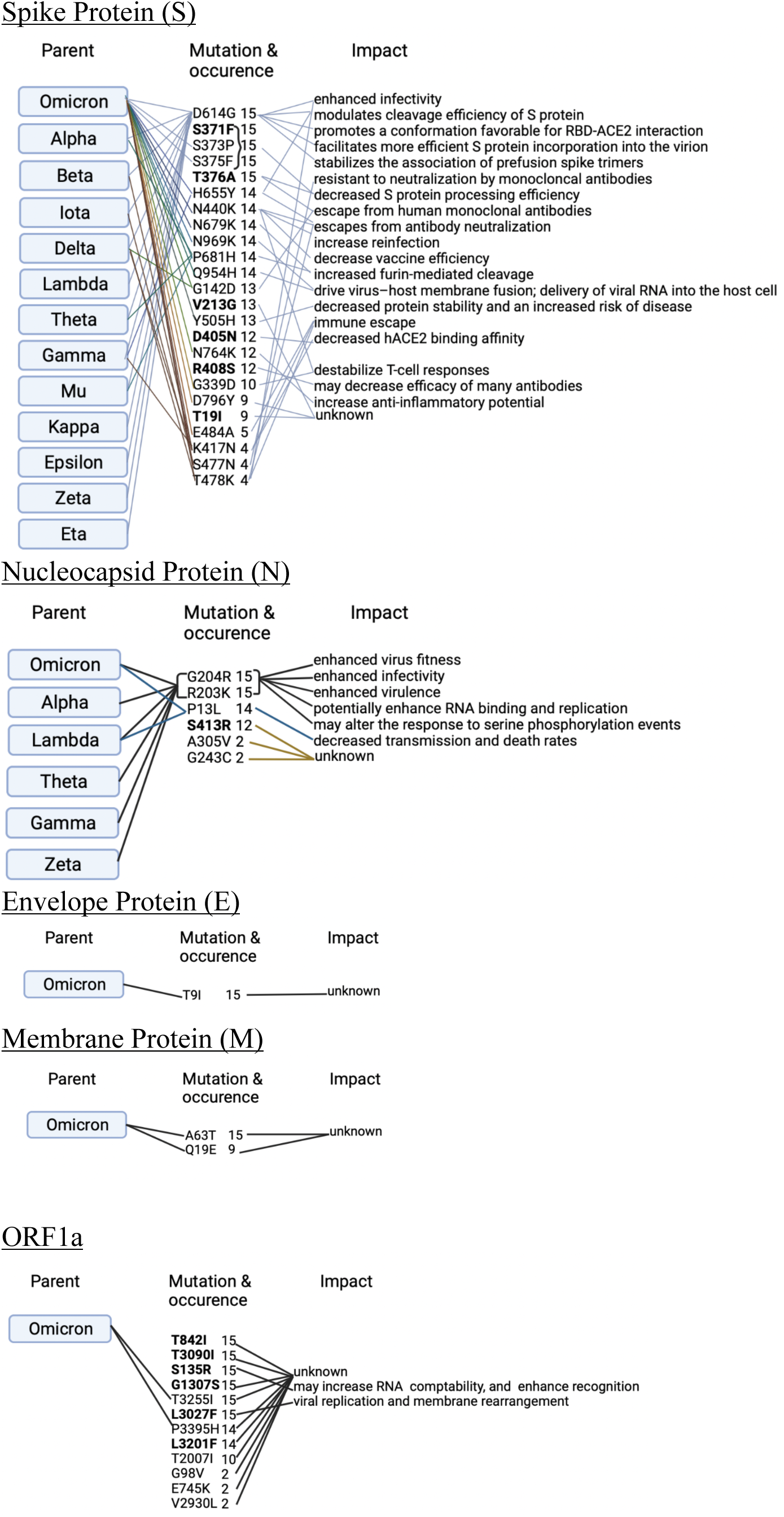
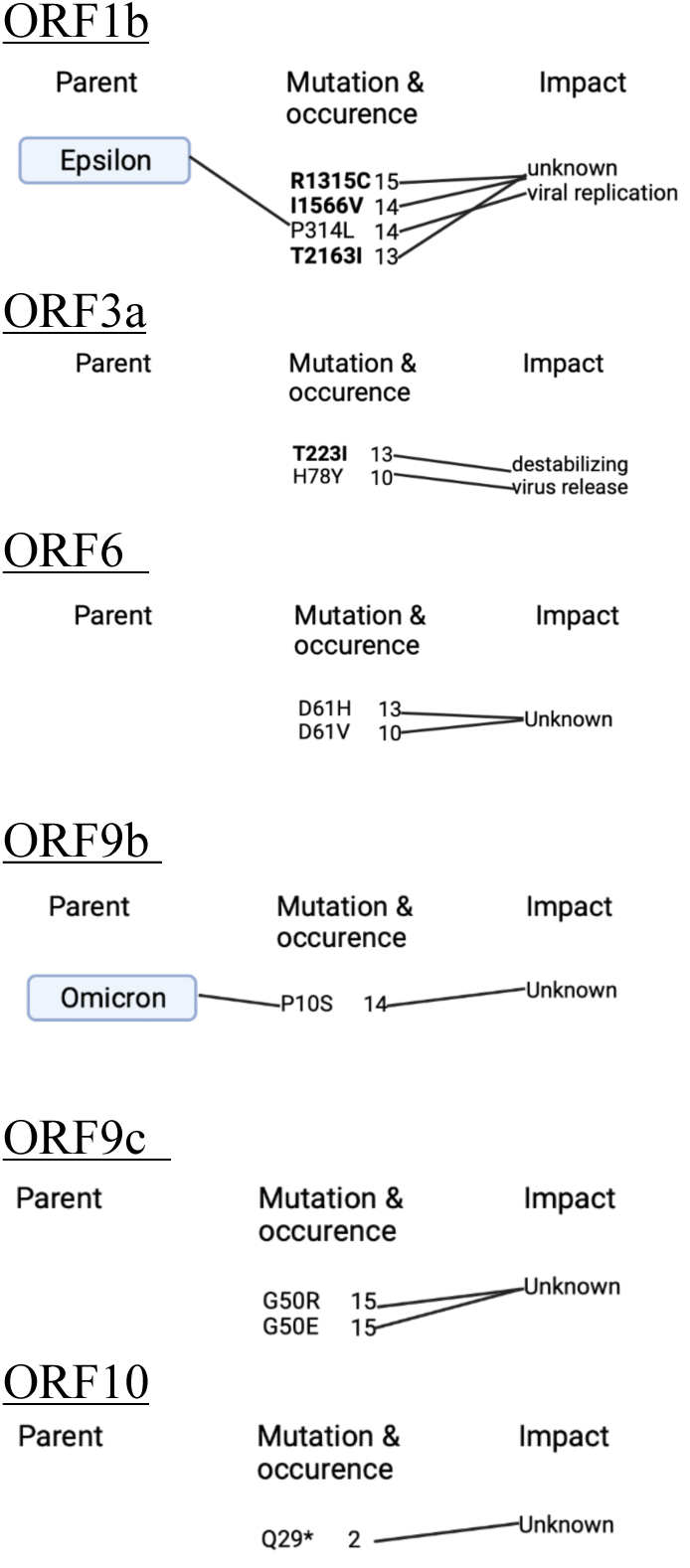
Amino acid mutations (occurring in at least 90% of the samples) in the wastewater for Omicron BA.2, parental origins and associated impacts. Mutations highlighted in **bold** were first observed in this variant and are subsequently observed in descendants Omicron BA.2.12.1, BA.4 and BA.5 as well; corresponding numbers indicate the frequency; mutations in parentheses indicate mutations that are known to occur together.

**Fig. S3.**
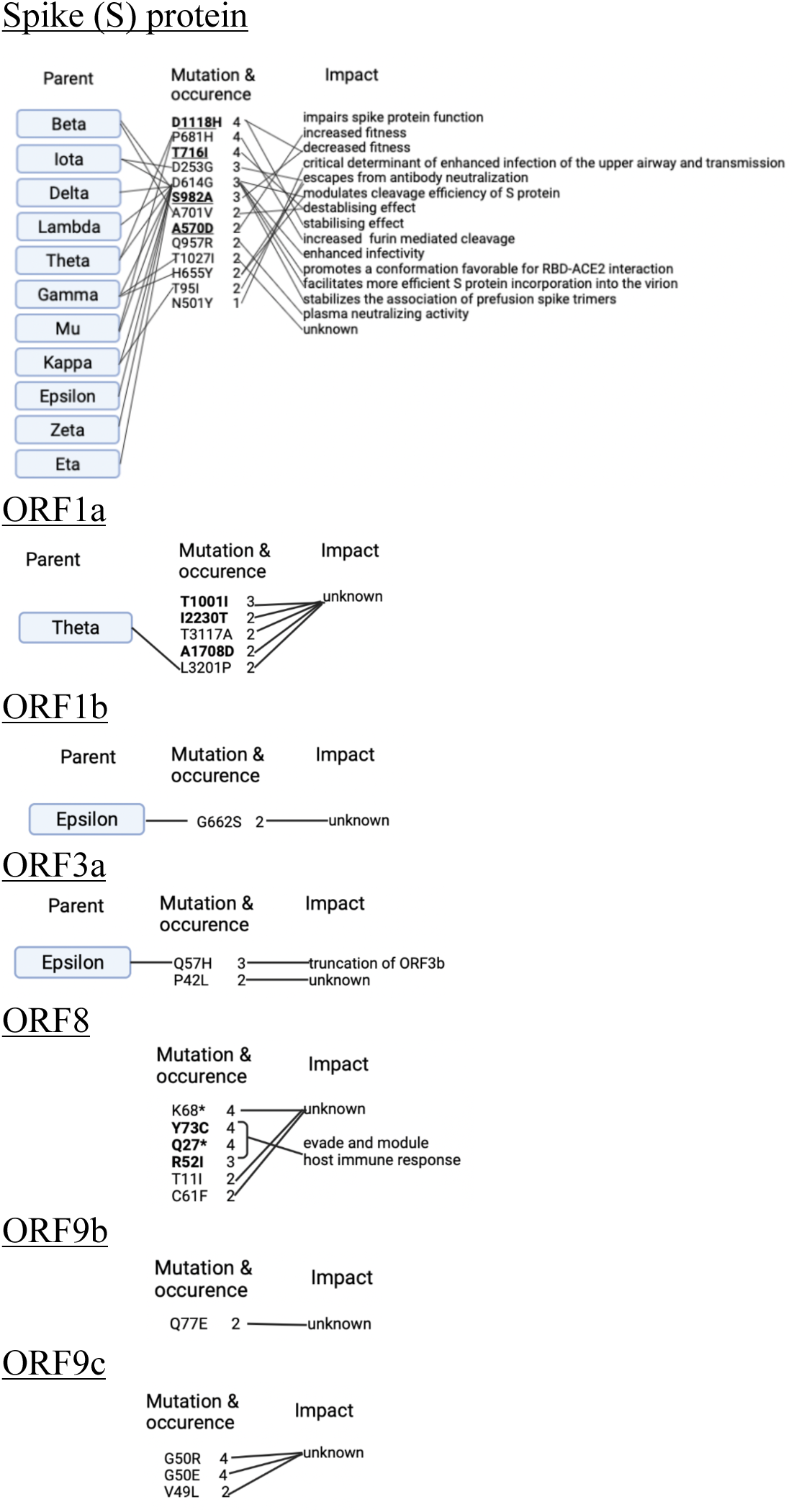
Amino acid mutations (occurring in at least 90% of the samples) in the wastewater for Alpha B.1.1.7, connections with other SARS-CoV-2 variants and associated impacts on functionality. Mutations highlighted in **<u>bold and underlined</u>** are unique to this variant; mutations highlighted in **bold only** were first observed in this variant and are subsequently observed in other descendants; corresponding numbers indicate the frequency.

**Fig. S4.**
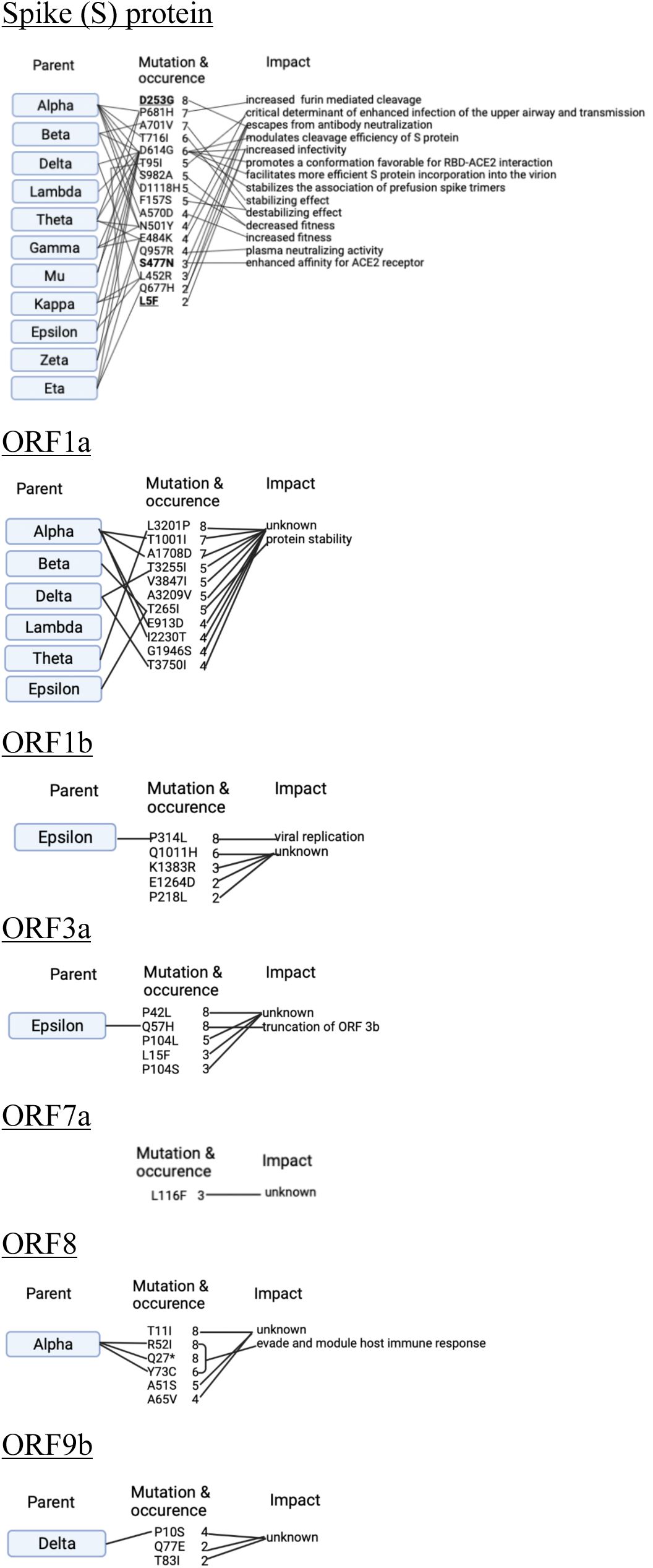
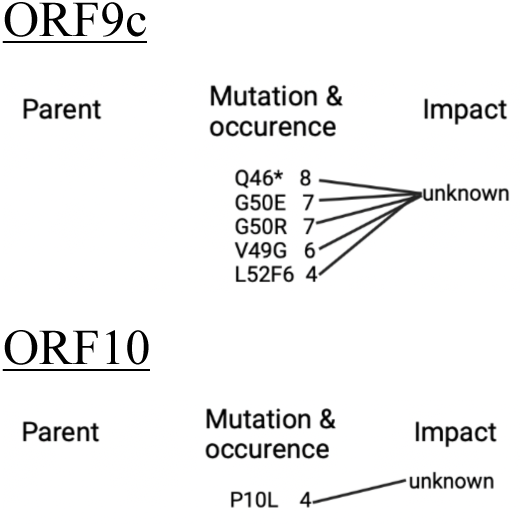
Amino acid mutations (occurring in at least 90% of the samples) in the wastewater for Iota B.1.526, connections with other SARS-CoV-2 variants and associated impacts on functionality. Mutations highlighted in **<u>bold and underlined</u>** are unique to this variant; mutations highlighted in **bold only** were first observed in this variant and are subsequently observed in other descendants; corresponding numbers indicate the frequency

**Fig. S5.**
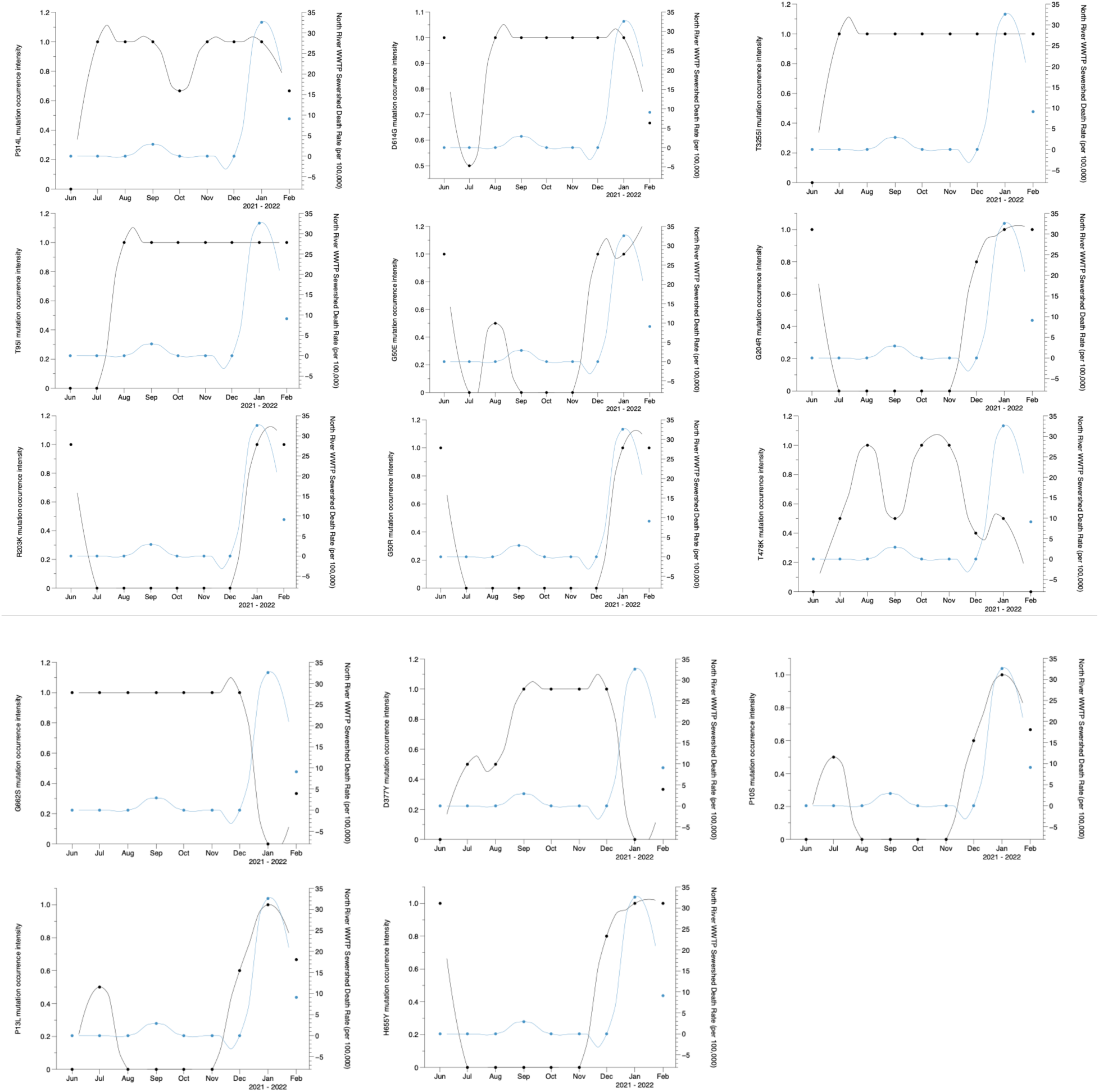
Amino acid mutation intensity (mutation frequency divided by number of samples and averaged over a month) in wastewater and COVID-19 related mortality rates (average death rate per 100,000 residents) in North River WWTP sewershed, New York City. The black solid line represents fitted curves for mutations, and blue solid line represents fitted curves for mortality rates calculated using the LOESS (locally estimated scatterplot smoothing) method.

**Fig. S6.**
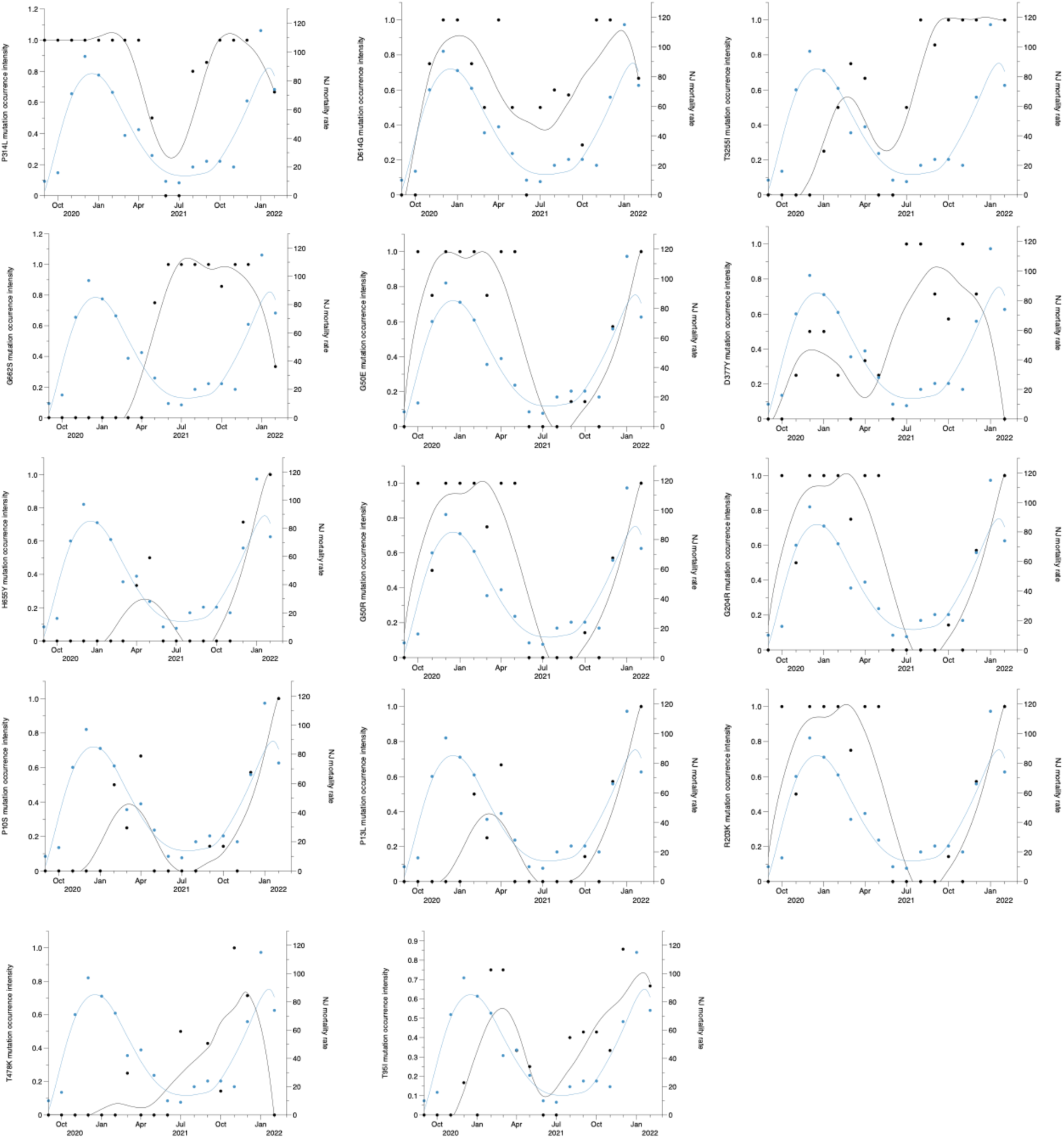
Amino acid mutation intensity (mutation frequency divided by number of samples and averaged over a month) in wastewater and COVID-19 related mortality rates (average death rate per 100,000 residents) in Little Ferry WWTP in Bergen County, New Jersey. The black solid line represents fitted curves for mutations, and blue solid line represents fitted curves for mortality rates calculated using the LOESS (locally estimated scatterplot smoothing) method.

**Fig. S7.**
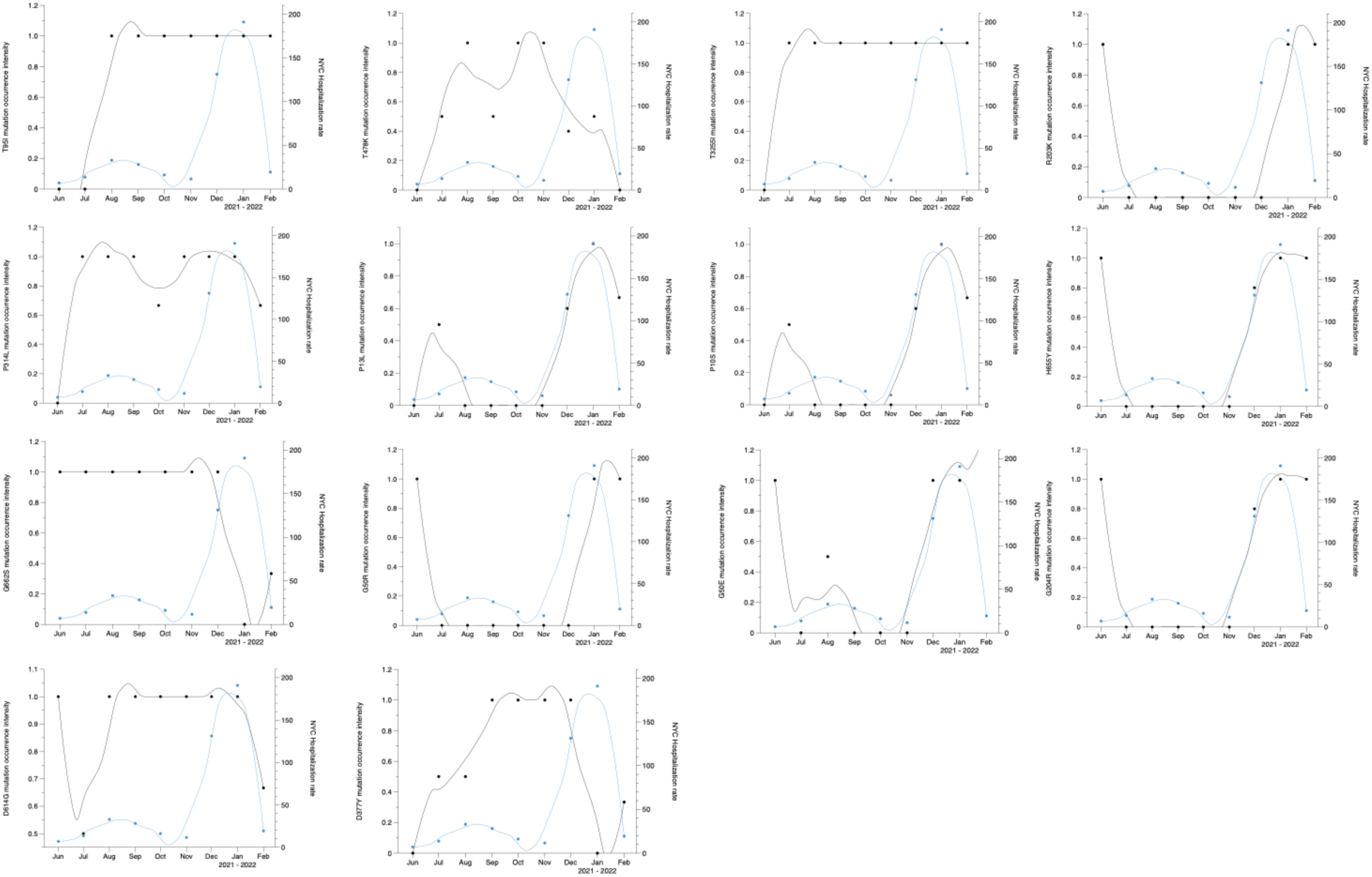
Amino acid mutation intensity (mutation frequency divided by number of samples and averaged over a month) in wastewater and COVID-19 related hospitalization rates (average number of laboratory confirmed cases who were ever hospitalized per month) in North River WWTP sewershed, New York City. The black solid line represents fitted curves for mutations, and blue solid line represents fitted curves for hospitalization rates calculated using the LOESS (locally estimated scatterplot smoothing) method.

**Fig. S8.**
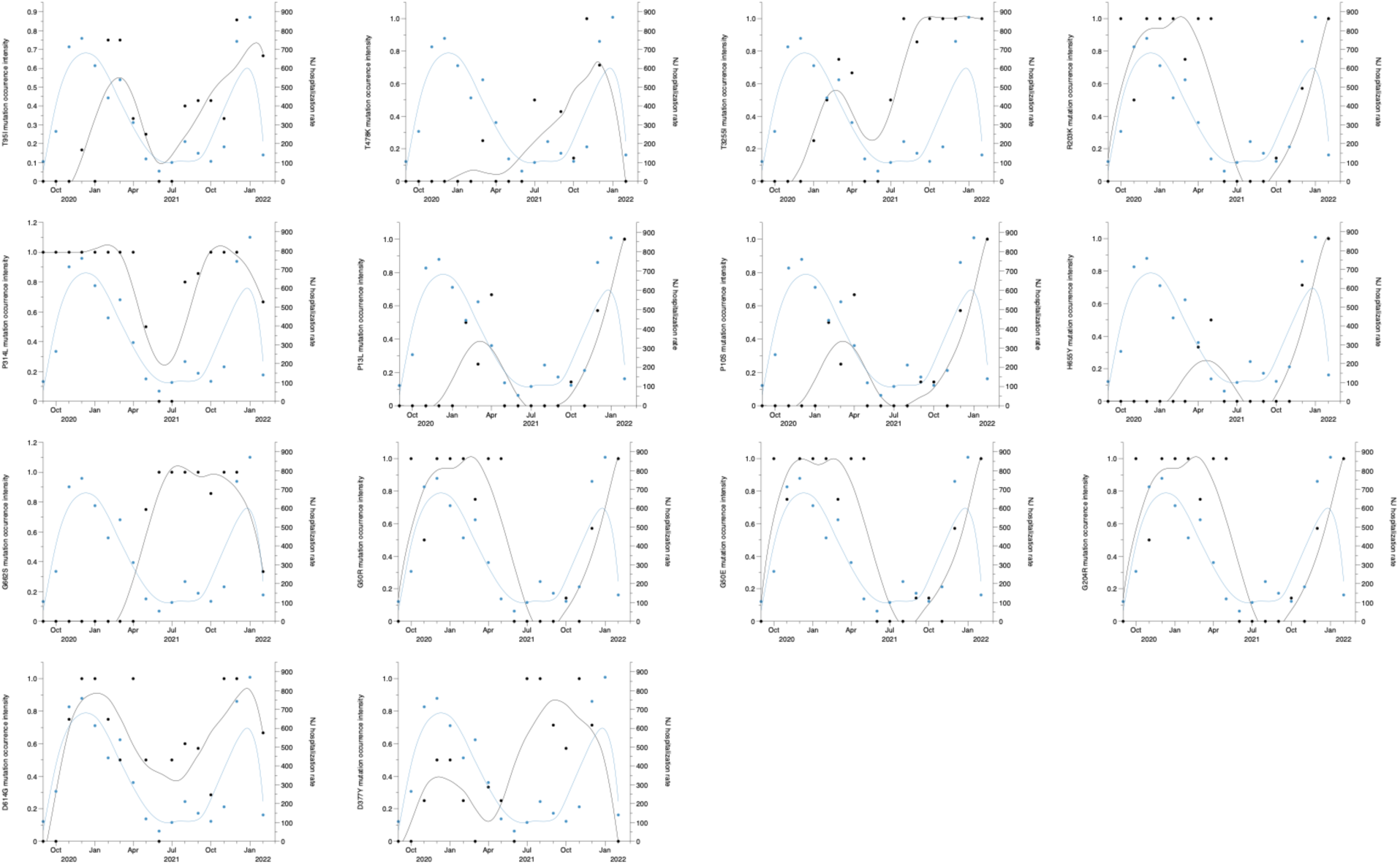
Amino acid mutation intensity (mutation frequency divided by number of samples and averaged over a month) in wastewater and COVID-19 related hospitalization rates (average number of laboratory confirmed cases who were ever hospitalized per month) in Little Ferry WWTP in Bergen County, New Jersey. The black solid line represents fitted curves for mutations, and blue solid line represents fitted curves for hospitalization rates calculated using the LOESS (locally estimated scatterplot smoothing) method.

**Fig. S9.**
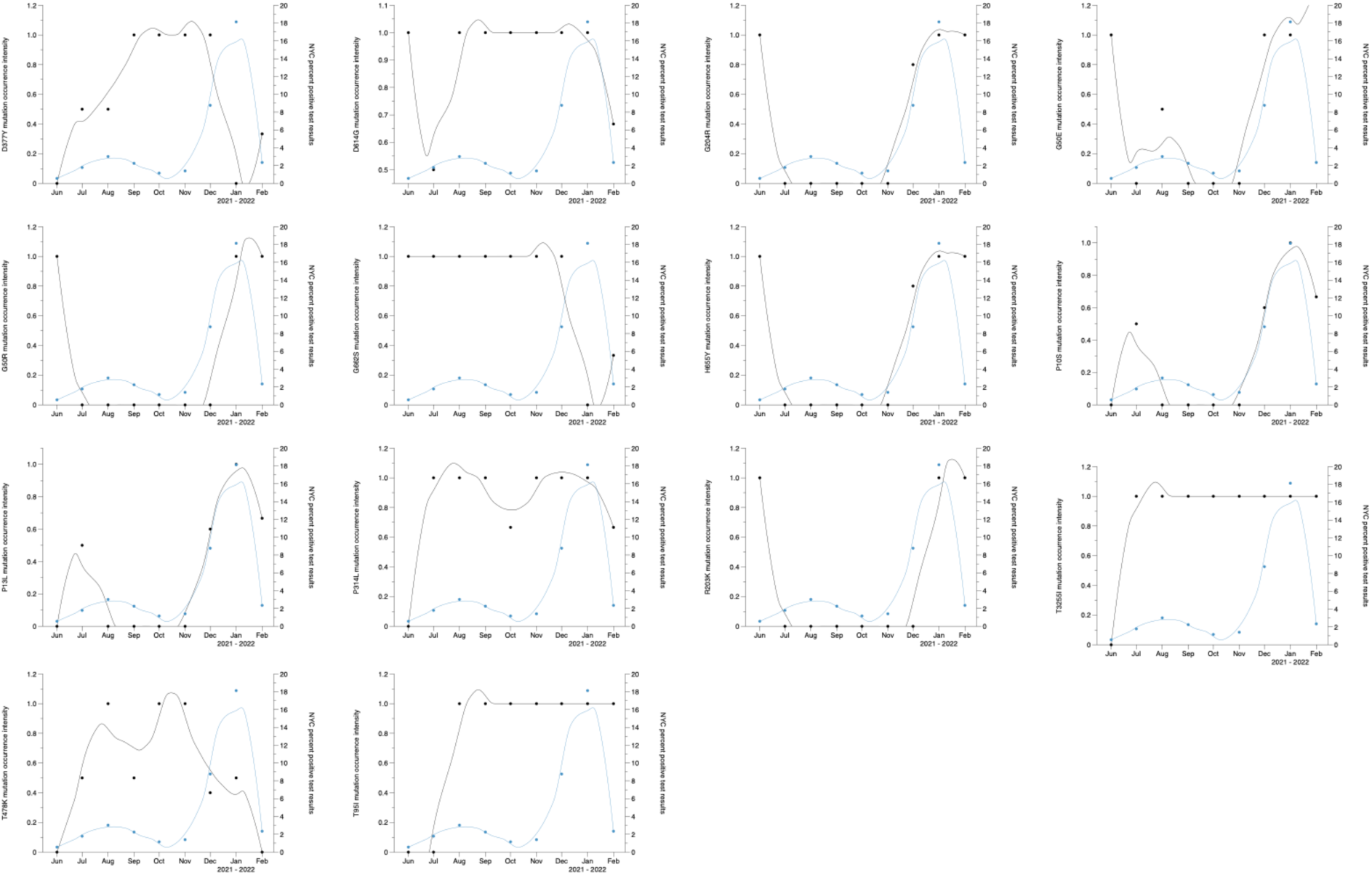
Amino acid mutation intensity (mutation frequency divided by number of samples and averaged over a month) in wastewater and COVID-19 monthly percent positivity PCR test results in North River WWTP sewershed, New York City. The black solid line represents fitted curves for mutations, and blue solid line represents fitted curves for percent positive rates calculated using the LOESS (locally estimated scatterplot smoothing) method.

**Fig. S10.**
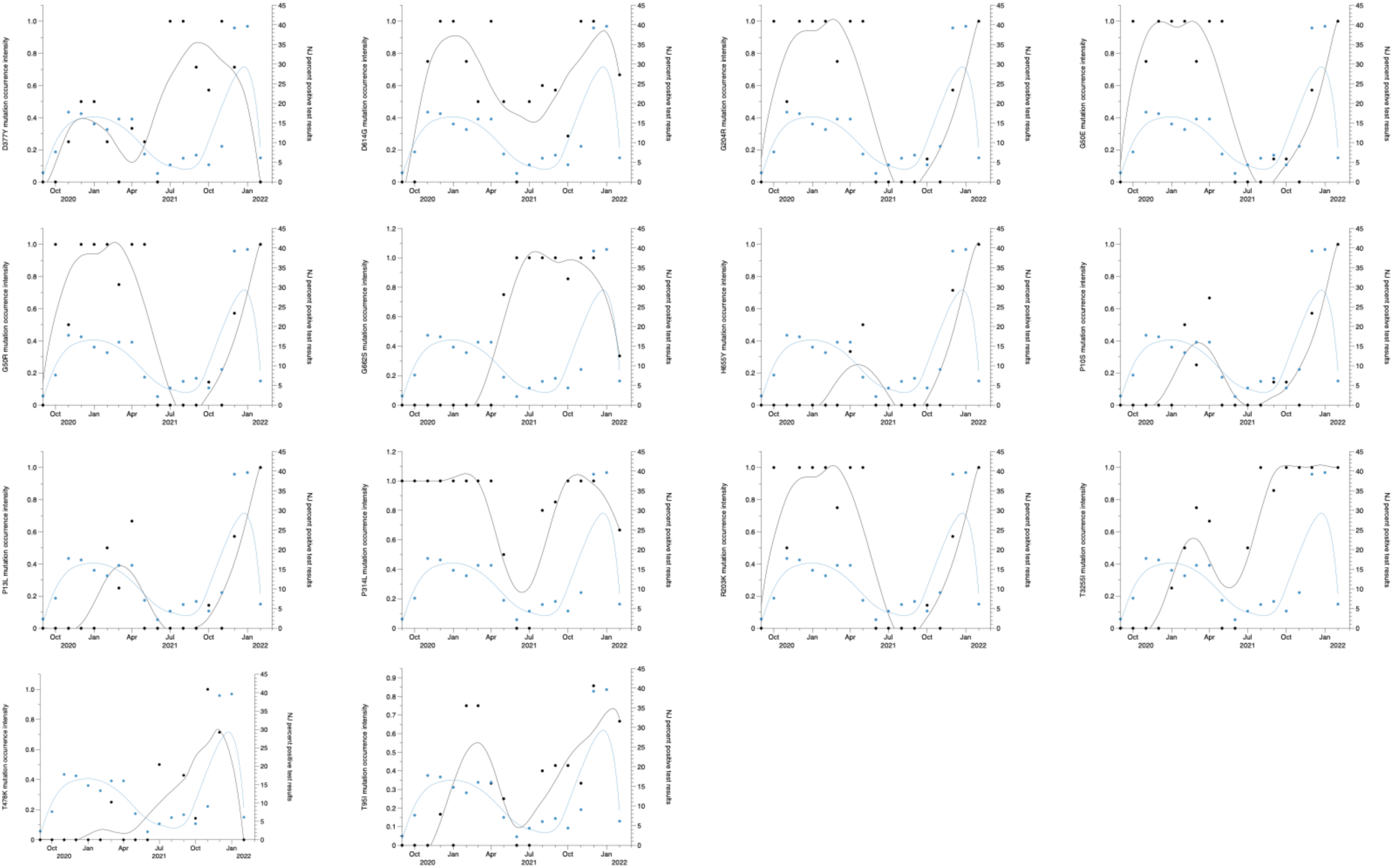
Amino acid mutation intensity (mutation frequency divided by number of samples and averaged over a month) in wastewater and COVID-19 monthly percent positivity PCR test results in Little Ferry WWTP in Bergen County, New Jersey. The black solid line represents fitted curves for mutations, and blue solid line represents fitted curves for percent positive rates calculated using the LOESS (locally estimated scatterplot smoothing) method.

**Fig. S11.**
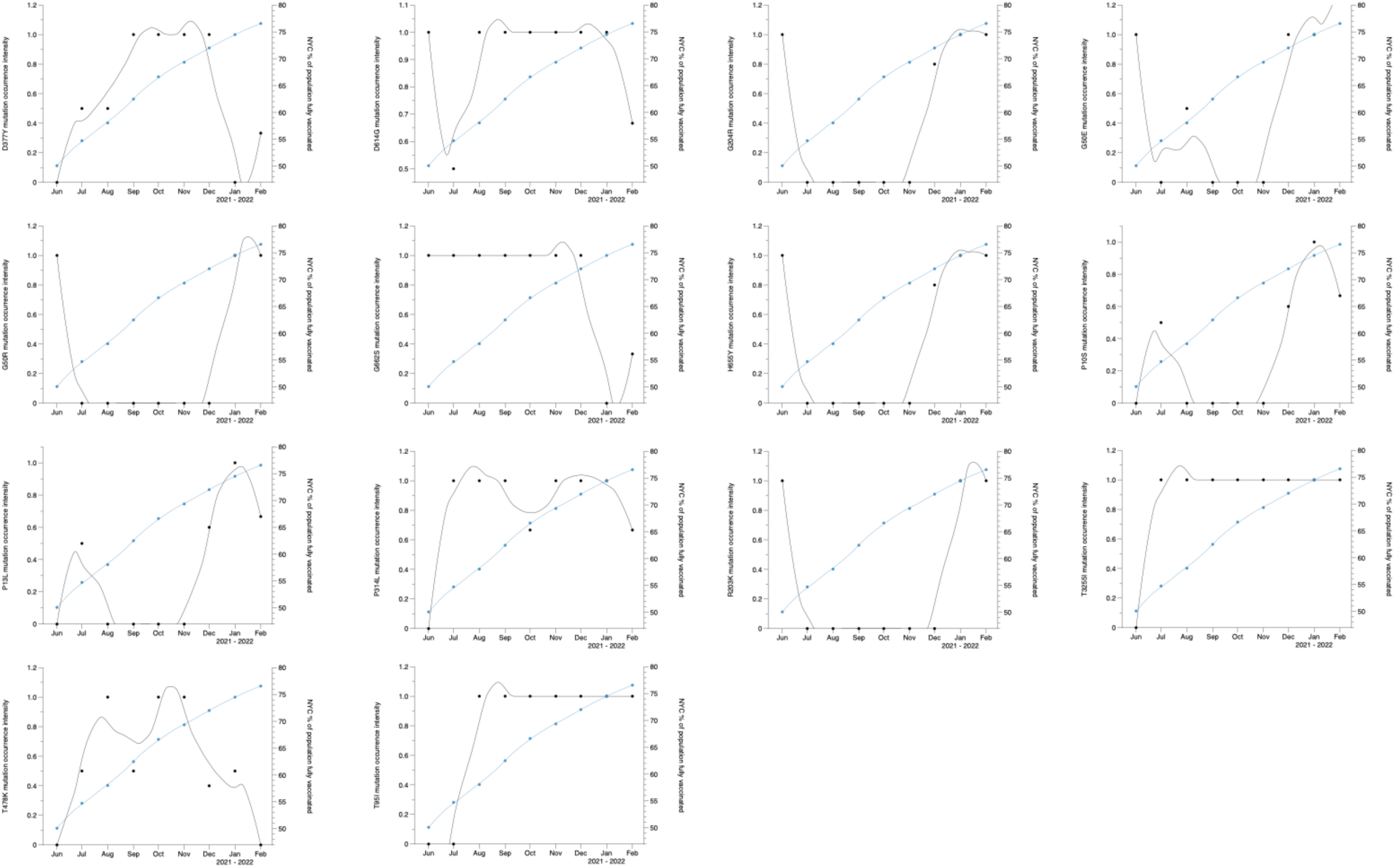
Amino acid mutation intensity (mutation frequency divided by number of samples and averaged over a month) in wastewater and % of population complete with COVID-19 vaccine courses in North River WWTP sewershed, New York City. The black solid line represents fitted curves for mutations, and blue solid line represents fitted curves for of population complete with COVID-19 vaccine courses calculated using the LOESS (locally estimated scatterplot smoothing) method.

**Fig. S12.**
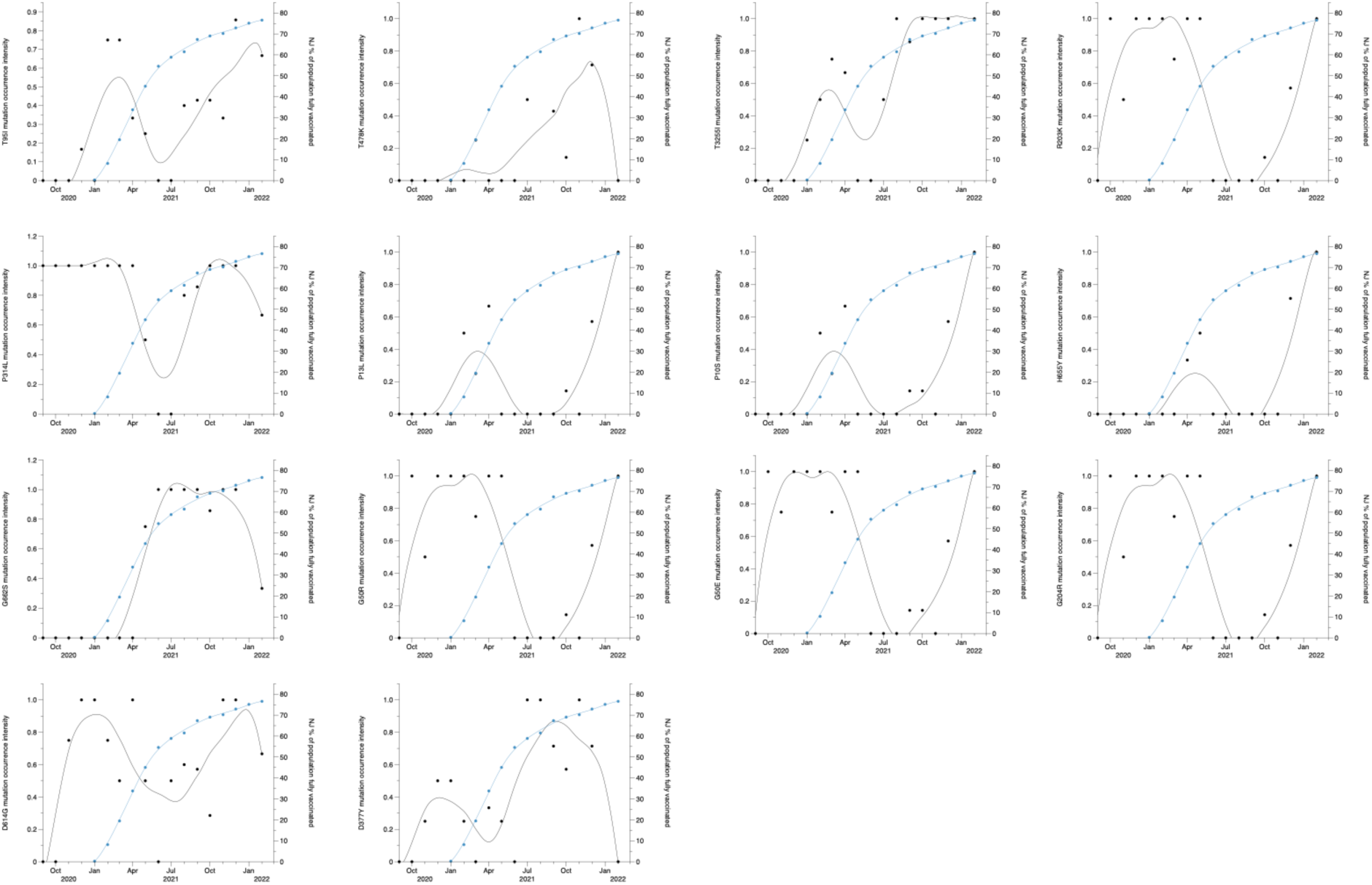
Amino acid mutation intensity (mutation frequency divided by number of samples and averaged over a month) in wastewater and % of population complete with COVID-19 vaccine courses in Little Ferry WWTP in Bergen County, New Jersey. The black solid line represents fitted curves for mutations, and blue solid line represents fitted curves for of population complete with COVID-19 vaccine courses calculated using the LOESS (locally estimated scatterplot smoothing) method.

